# Strategies to Mitigate COVID-19 Resurgence Assuming Immunity Waning: A Study for Karnataka, India

**DOI:** 10.1101/2021.05.26.21257836

**Authors:** Aniruddha Adiga, Siva Athreya, Bryan Lewis, Madhav V. Marathe, Nihesh Rathod, Rajesh Sundaresan, Samarth Swarup, Srinivasan Venkatramanan, Sarath Yasodharan

## Abstract

COVID-19 vaccination is being rolled out among the general population in India. Spatial heterogeneities exist in seroprevalence and active infections across India. Using a spatially explicit age-stratified model of Karnataka at the district level, we study three spatial vaccination allocation strategies under different vaccination capacities and a variety of non-pharmaceutical intervention (NPI) scenarios. The models are initialised using on-the-ground datasets that capture reported cases, seroprevalence estimates, seroreversion and vaccine rollout plans. The three vaccination strategies we consider are allocation in proportion to the district populations, allocation in inverse proportion to the seroprevalence estimates, and allocation in proportion to the case-incidence rates during a reference period.

The results suggest that the effectiveness of these strategies (in terms of cumulative cases at the end of a four-month horizon) are within 2% of each other, with allocation in proportion to population doing marginally better at the state level. The results suggest that the allocation schemes are robust and thus the focus should be on the easy to implement scheme based on population. Our immunity waning model predicts the possibility of a subsequent resurgence even under relatively strong NPIs. Finally, given a per-day vaccination capacity, our results suggest the level of NPIs needed for the healthcare infrastructure to handle a surge.

## 1 Introduction

India has witnessed a significant resurgence in the number of COVID-19 reported cases since the beginning of March 2021. The number of confirmed active cases which was 165,412 on 01 March 2021 increased thirteen-fold to 2,284,411 by 21 April 2021. The state of Karnataka, a large state in South India with a population of about 70 million (estimated for 2020) and an area of approximately 192,000 km^2^, also experienced this resurgence: from 5,945 confirmed active cases on 01 March 2021 to 196,236 confirmed active cases on 21 April 2021; the rate of positive tests increased from 0.68% to 15.87%, the number in intensive care units increased from 116 to 985, and the cumulative deaths increased from 12,343 to 13,885 during the same period [17, 18]. This resurgence has significantly stressed the healthcare system. Further, the resurgence has been heterogeneous across the districts of Karnataka, with Bengaluru Urban, Bidar, Kalaburagi, Udupi, Dakshina Kannada, and Tumakuru showing early resurgence.

India approved the use of two vaccines whose effectiveness from Phase 3 trials are now well-documented [6, 47]. To vaccinate 70%-90% of the population, Karnataka needs 98 – 126 million doses. This will likely take 6-24 months given current vaccine production levels in India (approximately 65 million doses per month during April 2021), production ramp-up plans, and all India requirements (Karnataka has about 1/20 of India’s population). The strategy has thus far prioritised (*i*) healthcare workers and front-line workers from 16 January 2021; (*ii*) those 60 years and above plus those 45 years and above with co-morbidities, from 01 March 2021, given that COVID-19 disproportionately affects older adults and individuals with comorbidities; and (*iii*) those 45 years and above from 01 April 2021. From 01 May 2021, all individuals 18 years and above became eligible for vaccination [35]. The challenges involved in vaccinating nearly a billion adults at the national level are outlined in [29].

Given the limited supply of vaccines, the eligibility of the entire adult population for vaccines, and Karnataka’s heterogeneous spread of COVID-19 across its districts, a study of *vaccination allocation policies across districts* is needed to design effective vaccination strategies. There are multiple possible criteria for vaccine allocation across districts – based on population, on seroprevalence, or on case-incidence rates. It is crucial to understand how these strategies compare in terms of minimising the number of cases. Assessing the effectiveness of these strategies is complicated due to many factors, including immunity waning, new variants, mobility, non-pharmaceutical interventions (NPIs)^1^, and delayed onset of vaccine effectiveness. To account for these factors and assess the overall impact of the vaccination strategies, we resort to a modeling study using a spatially explicit, age-stratified epidemiological model.

There are many district-level epidemiological models already available for Karnataka (See [1,3,25,40,43]). These models use time-series data on reported cases, confirmed active cases, tests conducted (some models), and deaths across the districts in the state. Age-stratified models have been used to understand vaccine allocation strategies for India around the time of the first wave [24]. While spatial allocations of COVID-19 vaccines have been studied in the recent literature [10, 13, 32], such a study has not been undertaken in the Indian context. We extend prior work by modelling the following additional factors.

- Karnataka conducted an extensive state-wide serological survey [4,5] during 02-16 September 2020 near the first peak. This information on the state of the pandemic at that time was used to initialise our model.
- Though more than 98% of the infected participants in a study seroconverted^2^ [48], it is well-documented that the antibody titres decay and the decay rate is robust across asymptomatics, mild symptomatics, age groups, sex, etc. [36] at approximately 25% reduction every two weeks. Antibody titres seem to be correlated with the severity of the infection [12, 22, 27, 33]. Data seems to suggest significant seroreversion among asymptomatics [30] who constitute a sizeable majority of the COVID-19 infected population in India [28]. We account for this seroreversion in our epidemiological model.
- As of 22 April 2021, about 6.7 million people, comprising healthcare workers, front-line workers, and people over 45 years of age, or just under 10% of the population of Karnataka, received at least one dose of vaccination in phases since 16 January 2021. We account for these vaccinations and their effectiveness in our model.
- There has been an increase in transmission during the resurgence phase, perhaps due to new variants in circulation. NPIs are being imposed for six weeks during April-June 2021 to break the chain of transmission [26]. We re-calibrate the contact rates to account for this resurgence and the NPIs.

By accounting for the above aspects in our epidemiological modelling, we study the following:

a. Effectiveness of three possible vaccination allocation strategies across districts: in proportion to the population, in inverse proportion to seroprevalence estimates, and in proportion to case-incidence rates;
b. Possibility of a subsequent resurgence in the latter half of 2021: under an immunity waning model along with various daily vaccination capacities and NPI scenarios; and
c. The target level of NPIs required: to ensure total caseload in a four-month horizon is within the healthcare infrastructure capacity given a daily vaccination budget.

The situation is fluid on the ground with significant ramping up of testing^3^, renewed telemedicine campaigns [20], and non-pharmaceutical interventions [26]. These will impact people’s behaviour, mobility patterns, and transmission dynamics. Nevertheless, we envisage that our study will provide epidemiological insights for better public health response and can serve as a framework that can be updated and expanded to a national scale based on data availability.

## 2. Methods: Data, Modelling, and Experiment Design

Karnataka has 30 administrative zones called districts. We treat all districts other than the capital Bengaluru Urban district as individual spatial units. Since the Bengaluru Urban district is heavily populated, we further subdivide it into nine units. This yields a total of 38 units which were also the units used in the state-wide serosurvey conducted in September 2021 [4, 5].

We use a spatial and age-stratified compartmental model to study the disease spread within Karnataka. There are five compartments: susceptible (S), exposed (E), infected (I), recovered/deceased (R) and vaccinated (V). The infected compartment includes both detected and undetected infections. The population is divided into sub-units, with each sub-unit comprising an (age-group, unit) pair, where a unit is one of the 38 units described above. The model keeps track of the numbers in the five compartments in every sub-unit and lets them evolve over time. The trajectory is obtained by time-discretisation and solutions to difference equations [45, 46], which we shall soon describe.

### Datasets

We use the following datasets in our model. The datasets used are portrayed in Figure 1 and summarised in Table 1.

- Population data is based on the official projected population data for 2020 [21]. The age distribution is based on a 2017 projection from [37].
- Age-stratified contact rates [38] modulate the number of contacts made between people of various age groups.
- Disease parameters (incubation period, mean recovery time) are based on [11, 23, 34].
- We leverage the unit-wise active infection and past infection data from the Karnataka-wide round-1 seroprevalence study [4, 5] to initialise our simulation on 11 October 2020.
- Time series of daily reported cases data and tests conducted in each unit are taken from the official daily bulletins (e.g., [18], summarised in [42]) starting from 11 October 2020. The data until 22 April 2021 is used for model calibration, and the data until 11 May 2021 is used for validation and uncertainty quantification.
- To move from reported cases to infections, we use the the unit-wise reported cases-to-infections ratio (CIR) estimated from the Karnataka round-1 seroprevalence study [4, 5]. Further, we use testing data from March 2021 to modify the CIR on a weekly basis. This is to account for significantly increased testing since March 2021. The testing data are taken from Karnataka’s daily COVID-19 bulletins.
- We also use the daily number of vaccinations administered in each unit and assume, for simplicity, a conservative 66% efficacy 14 days after one dose^4^.

**Figure 1:**
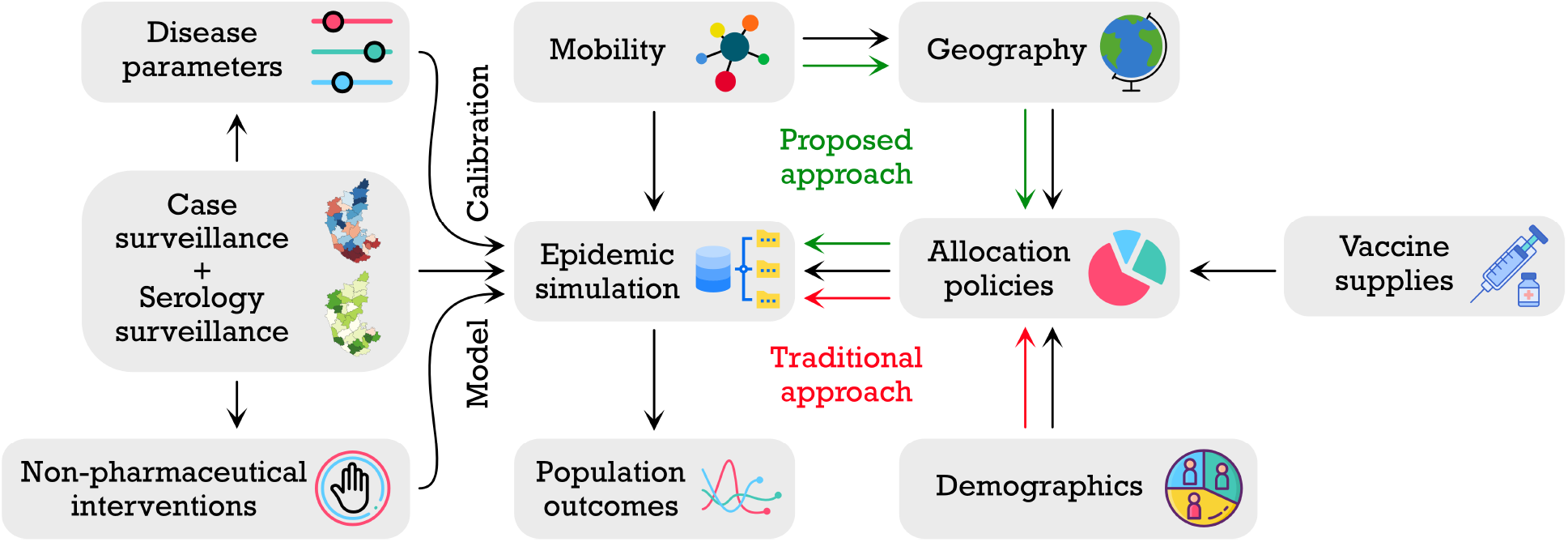
Datasets for our epidemic simulator. The epidemic simulator takes data from several sources. Disease parameters come from the current state of knowledge of how COVID-19 affects an infected individual. Case surveillance data include tests, reported cases, discharges, and deaths. Serosurveillance data comes from a state-wide survey done in September 2020 during the first peak. The timings and the duration of the NPIs are based on when they come into effect in the state. A mobility model captures the spread of infection in the units before the April 2021 resurgence. Karnataka’s actual geography enables better visualisation of the simulation outcomes. Demographic data for the units are taken from the 2011 census and extrapolated to 2020. Vaccine supply assumptions are based on historical vaccination data. Experimental vaccination allocation policies are based on these data elements. The epidemic simulator takes all these and generates outcomes for a comparative study.

**Table 1:** Summary of datasets used in the simulator, their brief descriptions, and references.

| Dataset | Description | Reference |
| --- | --- | --- |
| Population | The population of each unit of Karnataka (estimated for 2020). | [21] |
| Age distribution | The age distribution of the population in Karnataka. This is used to find out the population of each age group in each unit. | [37] |
| Age stratified contact rates | The number of contacts made by an individual of a given age group with another individual of another age group. This is used to modulate the contact rates based on age-groups. | [38] |
| Disease parameters | The mean incubation time in and the mean recovery time of COVID-19 infected individuals. These are used to model the spread of the disease. | [11, 23, 34] |
| Seroprevalence data | Unit-wise antibody (IgG) prevalence and active prevalence of COVID-19 from the Karnataka-wide round-1 serosurvey conducted during 02–16 September 2020. The round-1 data is used to initialise the simulator. We then use a Weibull model for immunity waning. | [4, 5] |
| Daily new reported cases | The time-series of unit-wise daily new reported cases since 11 October 2020. This is used to calibrate the contact rates. | [42] |
| Tests | The time-series of tests done in the state of Karnataka since 19 February 2021. This is used to modify the cases to infections ratio. | [42] |
| Vaccinations | The time-series of unit-wise vaccine doses administered since 16 January 2021. | [42] |

### Model

Let 𝒟 denote the set of units and let 𝒜 denote the set of age groups. A sub-unit (also referred to as a *patch*) of the population is identified with a given unit and an age group; let *u* = (*i, x*) ∈ 𝒟 × 𝒜 be a patch of the population. Let *N*_(*i,x*)_ denote the population of patch (*i, x*). Let us consider the evolution of the exposed population at *u*. People from patch *u* get exposed to the virus from the infected population in *u* as well as from the infected population in other patches *v* owing to mobility across units and interactions across age-groups. We model the impact of mobility on infection spread by keeping the population of the patches constant and by adjusting the within-unit and cross-unit contact rates based on a mobility matrix. Let Θ(*t*) denote the unit-wise travel matrix at time *t*, i.e., *θ*_*j,i*_(*t*) denotes the probability that an individual in unit *j* spends time in unit *i* at time *t*. Let *M* denote the matrix of age-stratified interaction rates, i.e., *M*_*x,y*_ is the number of contacts made by individuals of age group *x* with age group *y* on a typical day (time unit being one day). We also define *β*_*i*_(*t*), a piece-wise constant function, to be a contact rate modulating parameter at time *t*. Let *S*_(*i,x*)_(*t*), *E*_(*i,x*)_(*t*), *I*_(*i,x*)_(*t*), *R*_(*i,x*)_(*t*) and *V*_(*i,x*)_(*t*) denote the number of people in susceptible (S), exposed (E), infected (I), recovered/deceased (R), and vaccinated (V) states, respectively, in patch (*i, x*) at time *t*; see Figure 2. Assuming that a tagged individual in patch *u* = (*i, x*) comes in contact with a tagged individual in patch *v* = (*j, y*) at rate *β*_*i*_(*t*)*θ*_*j,i*_(*t*)*M*_*x,y*_*/N*_(*j,y*)_ effective contacts per unit time, the change in the exposed population at time *t* in patch *u* = (*i, x*), defined as Δ*E*_(*i,x*)_(*t*) = *E*_(*i,x*)_(*t* + 1) − *E*_(*i,x*)_(*t*), is given by

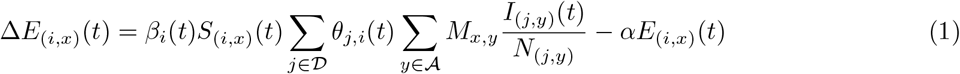

where *α* is the rate at which individuals move from the exposed state to the infected state (i.e., 1*/α* is the mean incubation period). Let *γ* denote the recovery rate, i.e., 1*/γ* is the mean time to recovery or death.

**Figure 2:**
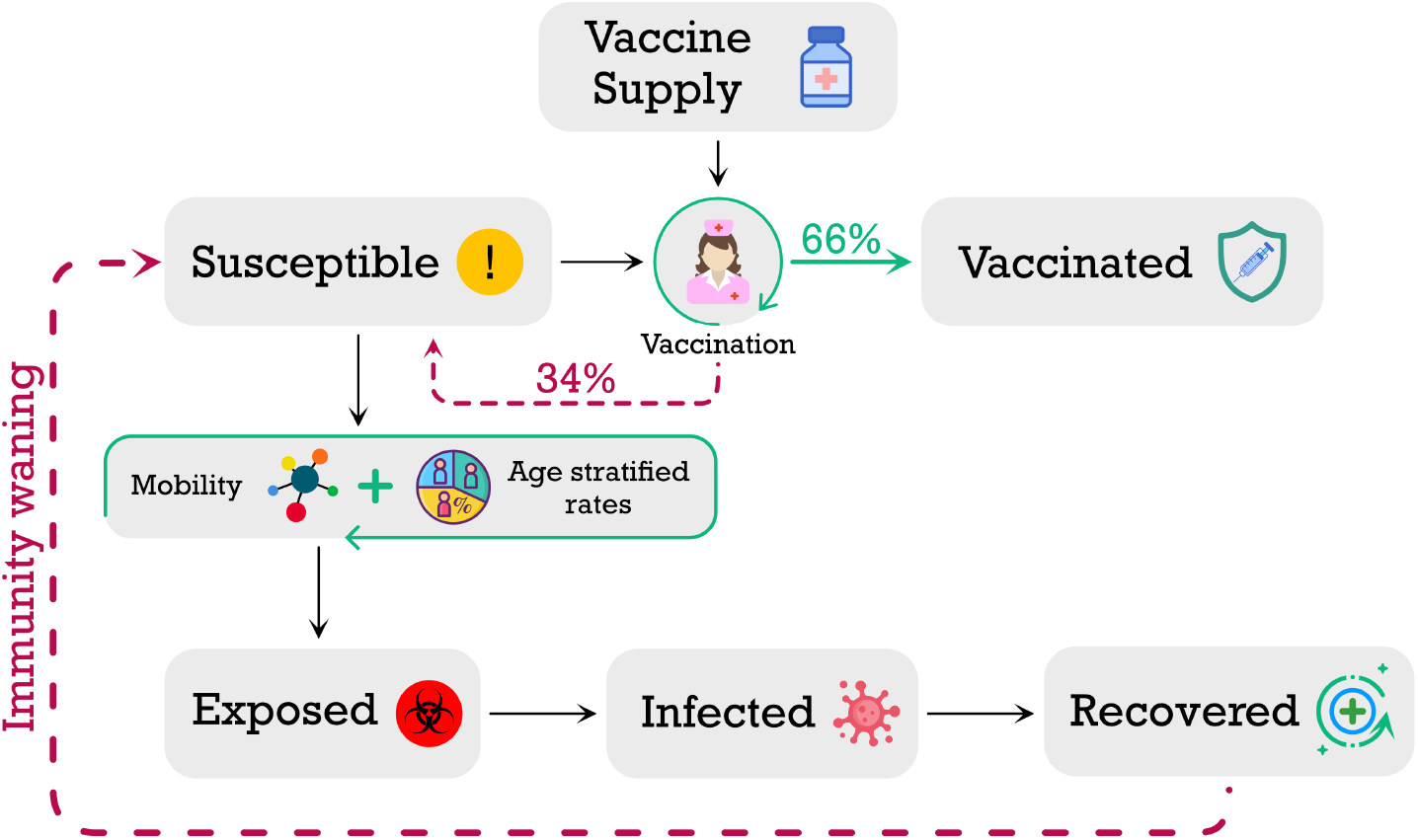
Compartments, disease progression, vaccination, and immunity waning (antibody decay) components of the model.

Then the changes in the variables *S, I*, and *R* for the patch (*i, x*) at time *t* are modelled as

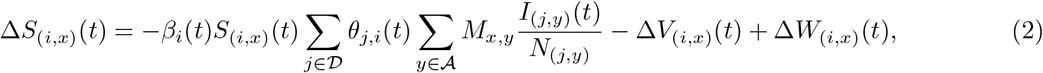

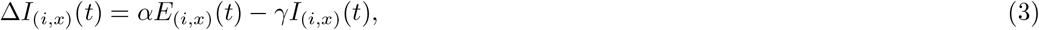

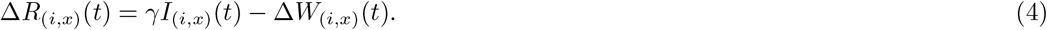

Additionally, *V*_(*i,x*)_(*t*) is incremented based on (for simplicity) the number of first-dose vaccines administered fourteen days prior to *t* in that unit, after taking into account the efficacy of the vaccine. This data is extracted from bulletins like [18] and is summarised in [42]. The 14-day delay accounts for the time for antibodies and immunity to develop. The increment to the vaccinated pool Δ*V*_(*i,x*)_(*t*) depends on the number vaccinated in age group *x* of district *i* and the effectiveness of the vaccine, which is conservatively set to 66%, the lower of the two effectiveness numbers reported in [6, 47] after two doses.

In addition to the disease progression dynamics (1)–(4), we also consider an agent-based immunity waning factor in our model. The feature, also known as seroreversion, is motivated by the observed reduction in seroprevalence in the densely populated areas of Mumbai [41]. Both reinfections from the original strain due to antibody waning and cross-strain infections are possible factors driving the increasing trend in the daily number of reported cases in Karnataka starting early March 2021. In our implementation of immunity waning and seroreversion, individuals in the recovered state are assumed to lose their immunity after a random amount of time which has the Weibull distribution with shape *κ* = 3.67 and scale *λ* = 120 days, a model inspired by that in [7] but with a faster median seroreversion period to account for the resurgence. We sample a duration from this Weibull distribution for each recovered individual and move that individual from the recovered state to the susceptible state after this duration. This is the Δ*W*_(*i,x*)_(*t*) in (2) and (4).

### Calibration

Certain parameters are kept fixed throughout the simulation duration. These are the mean incubation period 1*/α* (assumed to be 5.8 days) and the mean infectious period 1*/γ* (assumed to be 5 days). These are based on [11, 23, 34]. The piece-wise constant contact rate parameters *β*_*i*_() are tuned to minimise the per day squared error between the logarithm of daily-new-reported-cases time-series and that of the model (see Appendix 4.2). We aim to match the time-series for each unit by calibrating its contact rate. Since our simulator tracks infections, we multiply the reported cases by the estimated cases-to-infections ratio available from the Karnataka-wide round-1 seroprevalence study [4, 5] as the total infections that the simulator should track. To account for the increased testing since March 2021, we modify the CIR every week based on the ratio of the average number of daily tests done in a given week to the average number of daily tests done during 18–28 February 2021. Finally, we consider a 7-day moving average for the daily infected time-series to smooth the curves for tracking.

In summary, the following steps are performed during calibration:

- Start the simulator with susceptible, infected and recovered fractions matched to the round-1 seroprevalence data projected to 11 October 2020. Tune the contact rate parameters during 11–31 October 2020 to match the number of reported cases on 01 November 2020 within 10%.
- Tune the unit-wise contact rate parameters (one scalar per unit) during the period 01 November 2021 – 28 February 2021 to bring the per day squared error to within 0.1 for each unit. We use the identity matrix for mobility during this period, so the disease evolves independently in each unit. We update the contact rates in each iteration after evolving the time-series over the duration.
- Tune the unit-wise contact rate parameters during 01–15 March 2021 to match the number of model-reported cases on 15 March 2021 within 10% of the actual reported cases. Then tune the unit-wise contact rate parameters during 16 March – 07 April 2021 to minimise the per day squared error during this period. To account for the stochasticity due to a low number of reported cases in many units, we introduce moderate uniformly mixing mobility, i.e., Θ(*t*) = (1 *- ε*)*I* + *εJ*, where *I* denotes the identity matrix of size | 𝒟 |, *J* denotes the all-one matrix of size | 𝒟 |, and *ε* = 0.01.
- Repeat the above for the duration 08–22 April 2021 to arrive at the corresponding calibrated contact rates for this period. From this period onward, since the infections have been seeded, we go back to diagonal mobility, i.e., Θ(*t*) = *I*. If mobility data is available, then the mobility matrix Θ(*t*) can be appropriately set.
- For the projections, we redo the calibration for the period 08 April–01 May 2021 and hold the resultant contact rate parameters constant into the future.
- Finally, to account for the delay in sample collection and test outcomes, we delay the trajectory of reported cases by one week.

The immunity waning Weibull distribution is fixed to have shape *κ* = 3.67 and scale *λ* = 120 and is common across all units. This yields a median seroreversion time of 109 days.

### Validation

To validate the performance of our calibrated simulator, we use the tuned contact rates during 08–22 April 2021 and run the simulator with these contact rates during 23 April – 11 May 2021. We validate the simulator by comparing the simulator’s projections with the actual reported cases during 23 April – 11 May 2021. To quantify the uncertainty in our predictions, we compute the root mean squared error between the projected number of cases and the actual reported cases between 23 April – 11 May 2021.

### Experiment design

We use our simulator to estimate the number of reported cases for each unit in Karnataka until 31 December 2021. However, we caution that we do not have uncertainty quantifications for the longer-term projections (beyond two weeks). The experiments are described below.

We first consider two sets of scenarios to study the effect of non-pharmaceutical interventions (NPIs). These are the following.

- The first set is the setting of No-NPI. Here the mobility matrix during 02 May 2021 – 31 December 2021 is assumed to be Θ(*t*) = *I*, i.e., the same as that of the period 01 April 2021 – 01 May 2021 (violet extrapolation in Figure 3).
- The second set consists of three scenarios with varying levels of NPI during 02 May 2021 – 31 December 2021:

I. 1/3-NPI with mobility 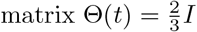
II. 1/2-NPI with mobility 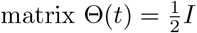
III. 2/3-NPI with mobility 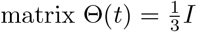

**Figure 3:**
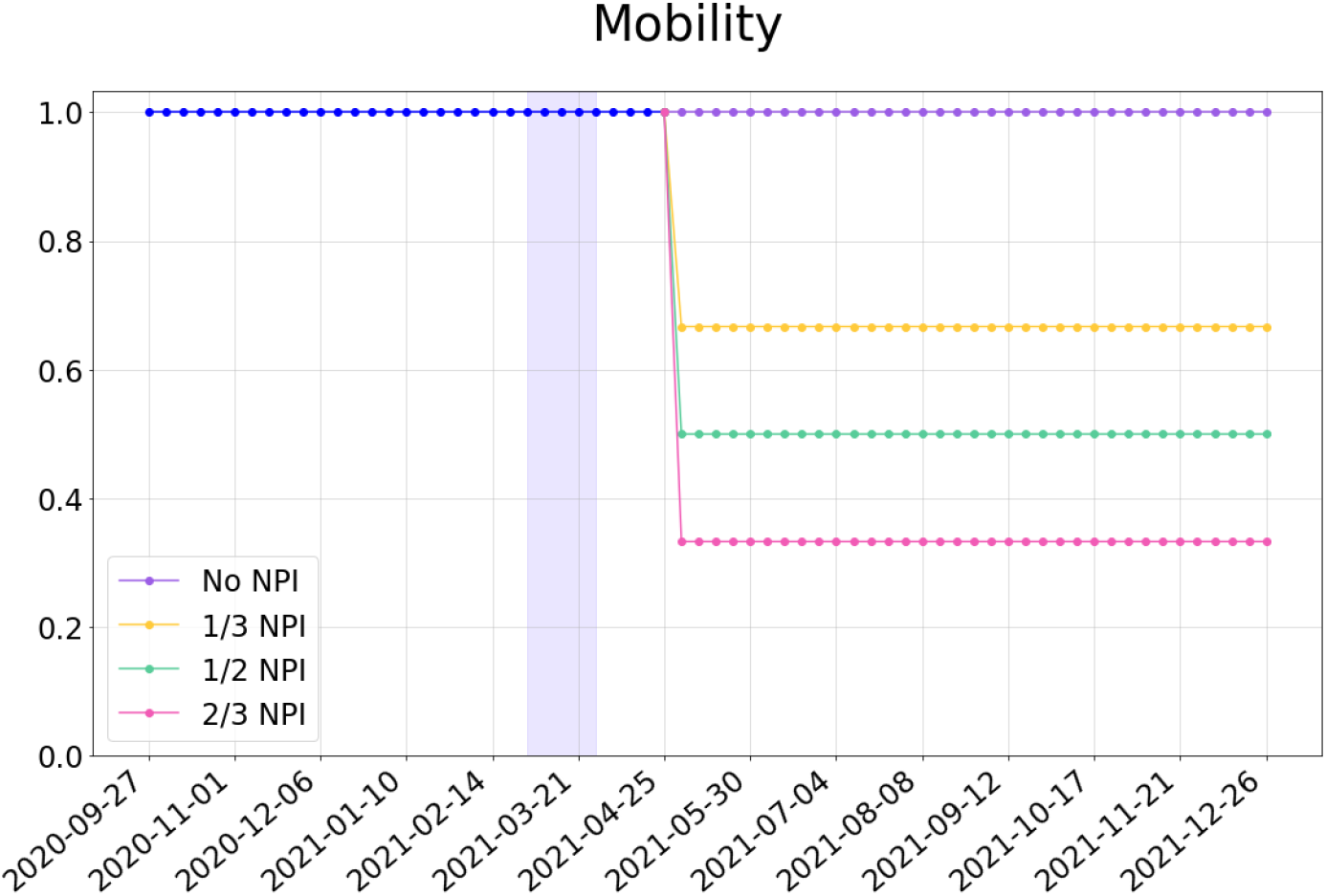
Mobility scenarios considered for the simulation experiment design. The purple band indicates a period when the mobility matrix is (1 *- ε*)*I* + *εJ*. The mobility is *I* at all other times.

The above scenarios are depicted in Figure 3. Note that Θ(*t*) = *αI* implies that the number *α* is the reduction in the effective contact rate from a reference level of 1. We choose to study three different levels to account for uncertainties in the actual level of NPI and compliance.

We also design three vaccination policies. Starting from 23 April 2021, vaccines are allocated to units in proportion to:

- The population of the units;
- The inverse of the model-predicted seroprevalence on 18 February 2021; and
- The case-incidence rates during 01–15 April 2021.

We have chosen 18 February 2021 as the reference date for the inverse-seroprevalence strategy to match the second seroprevalence study, which is in progress. For each policy above, starting from 23 April 2021, we assume that the daily budget of available vaccine doses is 167000 (which is the average daily number of first doses of vaccines administered in the state of Karnataka during 01–15 April 2021). Until 22 April 2021, we use the actual number of the first dose of the vaccine administered; this data is available in [16]. Since the vaccination drive started with healthcare workers and subsequently extended to people above 60, 45 and then 18 years of age, we assume that the vaccines allocated to a unit get equally distributed among the age groups of 18+ during 16 January 2021 – 15 March 2021 (healthcare workers), equally distributed among the age groups of 40+ during 16 March 2021 – 30 April 2021, and equally distributed among the age groups of 18+ after 01 May 2021, respectively. Even though India has a significantly younger population, in our simulator, the distribution of vaccines across age groups has a desirable natural bias towards the elderly, who are more susceptible to COVID-19.

Finally, we also study two decreased and seven increased daily vaccination capacities where the 167000 is replaced by 100000, 133000, 200000, 233000, 267000, …, 400000 doses per day (first dose). See Table 2.

**Table 2:** The projected number of cumulative reported cases in the state of Karnataka on 31 August 2021 under the four NPI intensities and various daily budgets of available vaccine doses. Vaccinations are assumed to be allocated in proportion to the population of each unit.

| Daily budget | No-NPI | 1/3-NPI | 1/2-NPI | 2/3-NPI |
| --- | --- | --- | --- | --- |
| 100000 | 3266600 | 2278100 | 1815200 | 1490700 |
| 133000 | 3206300 | 2249400 | 1803000 | 1487500 |
| 167000 | 3147900 | 2221800 | 1790400 | 1484600 |
| 200000 | 3094100 | 2196100 | 1779400 | 1480900 |
| 233000 | 3043300 | 2172400 | 1769400 | 1477400 |
| 267000 | 2993600 | 2148800 | 1758400 | 1474200 |
| 300000 | 2948300 | 2127600 | 1749200 | 1470900 |
| 333000 | 2905200 | 2107200 | 1740000 | 1468000 |
| 367000 | 2863000 | 2087000 | 1730900 | 1465500 |
| 400000 | 2824200 | 2069100 | 1722400 | 1462800 |

The outcomes of our experimental study are summarised using:

- The variation in the occurrence of the peak of daily reported cases across the units of Karnataka under various NPIs;
- The short-term and long-term cumulative number of reported cases in the units of Karnataka under the three vaccination policies; and
- The reduction in the number of projected cases with increase in the daily budget of available vaccine doses for various NPIs.

The outcomes will quantify the public health benefit of NPIs over No-NPI, study the effectiveness of the three vaccination policies, and highlight the limitations/benefits of reduced/increased vaccine allocations across various levels of NPIs.

## 3. Results

### Validation and uncertainty quantification

The prediction errors in the simulator’s projections for the entire state of Karnataka are visualised in Figure 4. The projections are in blue and should be compared with the actual reported cases in red on the right-hand side of the vertical line marking 22 April 2021. Similar validation plots for the individual units can be found in Figure 13. For the state of Karnataka, we find that the root mean squared error between the projected number of cases and the reported number of cases is about 9%. The percentage error for each unit is visualised in Figure 5. While the short-term prediction errors for more than half of the units are within 20%, the error for some units such as Raichur and Vijayapura are close to 50%. Longer-term prediction errors will only be larger. Further, there is a steep rise in the prediction errors for the units Chikkaballapur, Kodagu, Kolar, Vijayapura and Raichur.

**Figure 4:**
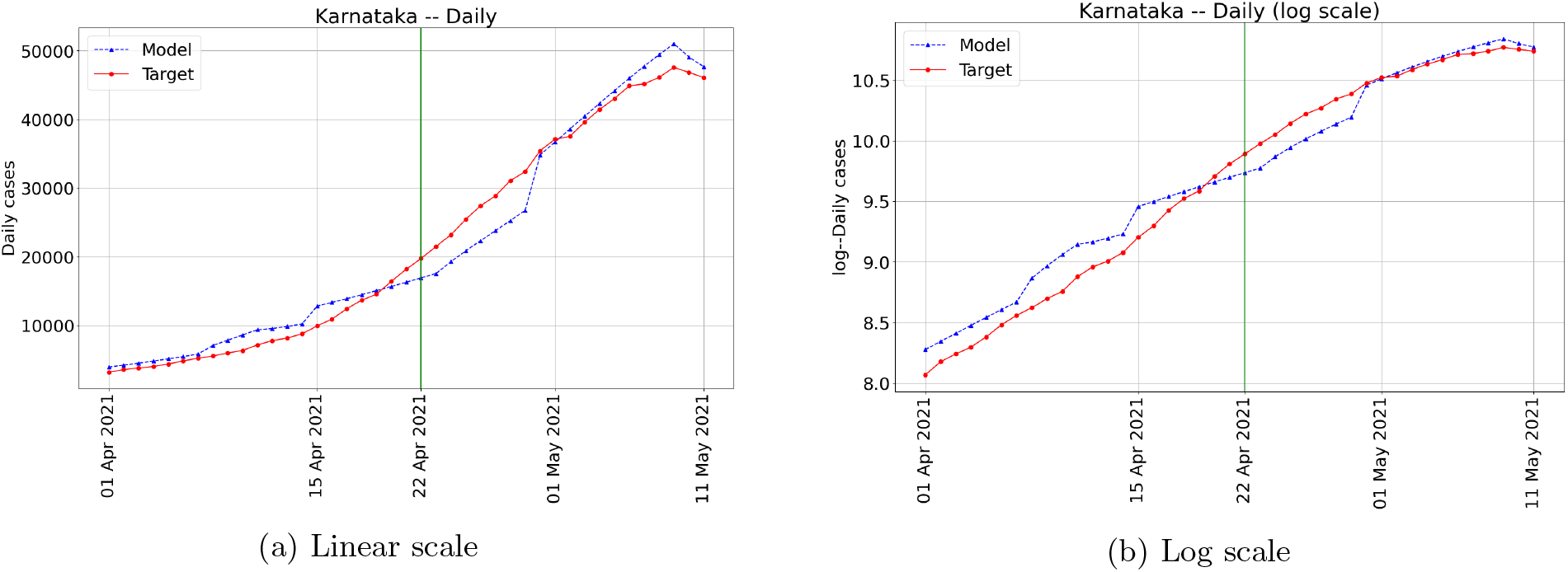
Validation plots for the state of Karnataka. The vertical line in both plots separates the calibration and the validation periods. The root mean squared error during the validation period is at most 9% for the state of Karnataka.

**Figure 5:**
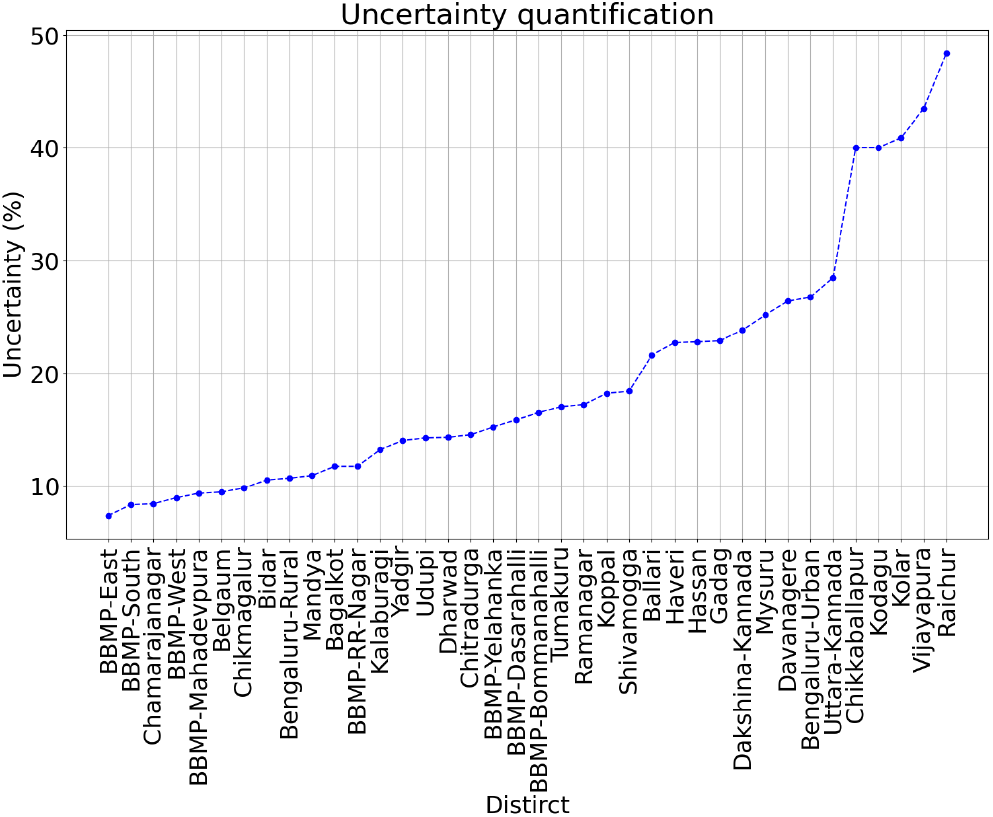
Uncertainty quantification for the validation period in each unit of Karnataka. More than one-half of the units have a root mean squared error of less than 20%. Five districts have errors between 40-50% which are due to the sudden change in trends in the reported cases not captured by the model.

### Future projections and peaks

Figure 6 depicts the daily and the cumulative number of reported cases in Karnataka until 31 December 2021 under various NPIs. In these plots, we assume that a daily budget of 167000 vaccine doses is available and distributed in proportion to each unit’s population. Note that these are long term projections and should be interpreted with caution.

**Figure 6:**
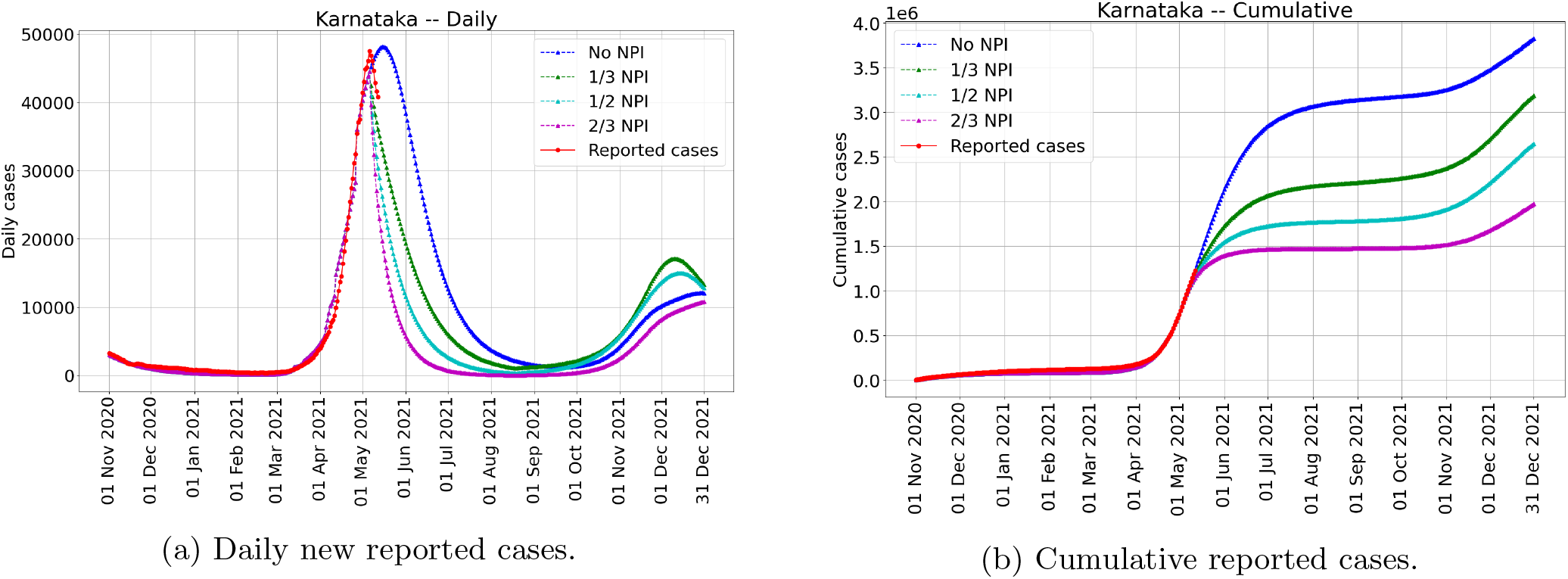
The projected number of new infections for the state of Karnataka under various NPIs. These projections are for the vaccination policy that allocates 167000 vaccine doses per day proportional to each unit’s population. Uncertainty bands are not indicated to reduce clutter. Recall the 9% root mean squared error for a two-week ahead projection. The left figure is for the daily new reported cases, and the right figure provides the cumulative reported cases.

The units are classified as E1 *<* E2 *<* M1 *<* M2 *<* L1 *<* L2 with E standing for early peaks (May 2021), M standing for mid-range peaks (June 2021), and L standing for late peaks (July 2021 and after). E1 and E2 stand for the first and second half of May 2021, respectively, with similar interpretations for M1, M2, and L1, L2.

#### Peaks – The counter-factual case of No-NPI

Under the counter-factual No-NPI scenario, the state-level peak may have occurred during mid-May 2021. For the unit-wise daily and the cumulative number of reported cases, see Figure 11. The model suggests that the units would have peaked at different times. Most of the Bengaluru units are in the E category (either E1 or E2) and Kalaburagi, Bidar, Kodagu, and Tumakuru. Except for Kodagu, these units saw an early resurgence. Kodagu saw a steep rise in infections, which suggests an early peak. Bagalkote, Chitradurga, Haveri, Dharwad and Gadag are in the L category. See the top two subfigures in Figure 9 for the No-NPI setting. The left-hand side figures present the E-M-L categorisation, while the right-hand side figures present a spatial view of this categorisation.

#### Peaks – The various NPIs

Under 1/3-NPI, the model predicts that only about a third of the units will have peaks after mid-May 2021 (E2 and beyond). Under 1/2-NPI and 2/3-NPI, the model predicts that cases reduce from as early as mid-May 2021 in every unit and also in the state.

#### Projected cumulative cases on 31 August 2021 – A comparison

The projected cumulative reported cases at the end of August 2021 seem to have significant reduction due to these NPI assumptions, see Figure 6 and Tables 4–7. These assume 167000 vaccination doses per day. Compared to No-NPI, on 31 August 2021, the model projects that 1/3-NPI may reduce the (cumulative reported) cases by roughly 926,000, 1/2-NPI may reduce it by another 431,000 cases, and 2/3-NPI may reduce it by another 306,000 cases. This order of magnitude reduction appears robust across all three vaccination policies, which we discuss next.

**Table 3:** Reduction in the number of cumulative projected cases on 31 August 2021 under various NPI intensities and daily vaccination budgets. Vaccinations are assumed to be allocated in proportion to the population of each unit.

| Daily budget | No-NPI | 1/3-NPI | 1/2-NPI | 2/3-NPI |
| --- | --- | --- | --- | --- |
| 100000 | Reference | 30.3% | 44.4% | 54.4% |
| 133000 | 1.8% | 31.1% | 44.8% | 54.5% |
| 167000 | 3.6% | 32.0% | 45.2% | 54.6% |
| 200000 | 5.3% | 32.8% | 45.5% | 54.7% |
| 233000 | 6.8% | 33.5% | 45.8% | 54.8% |
| 267000 | 8.4% | 34.2% | 46.2% | 54.9% |
| 300000 | 9.7% | 34.9% | 46.5% | 55.0% |
| 333000 | 11.1% | 35.5% | 46.7% | 55.1% |
| 367000 | 12.4% | 36.1% | 47.0% | 55.1% |
| 400000 | 13.5% | 36.7% | 47.3% | 55.2% |

**Table 4:**
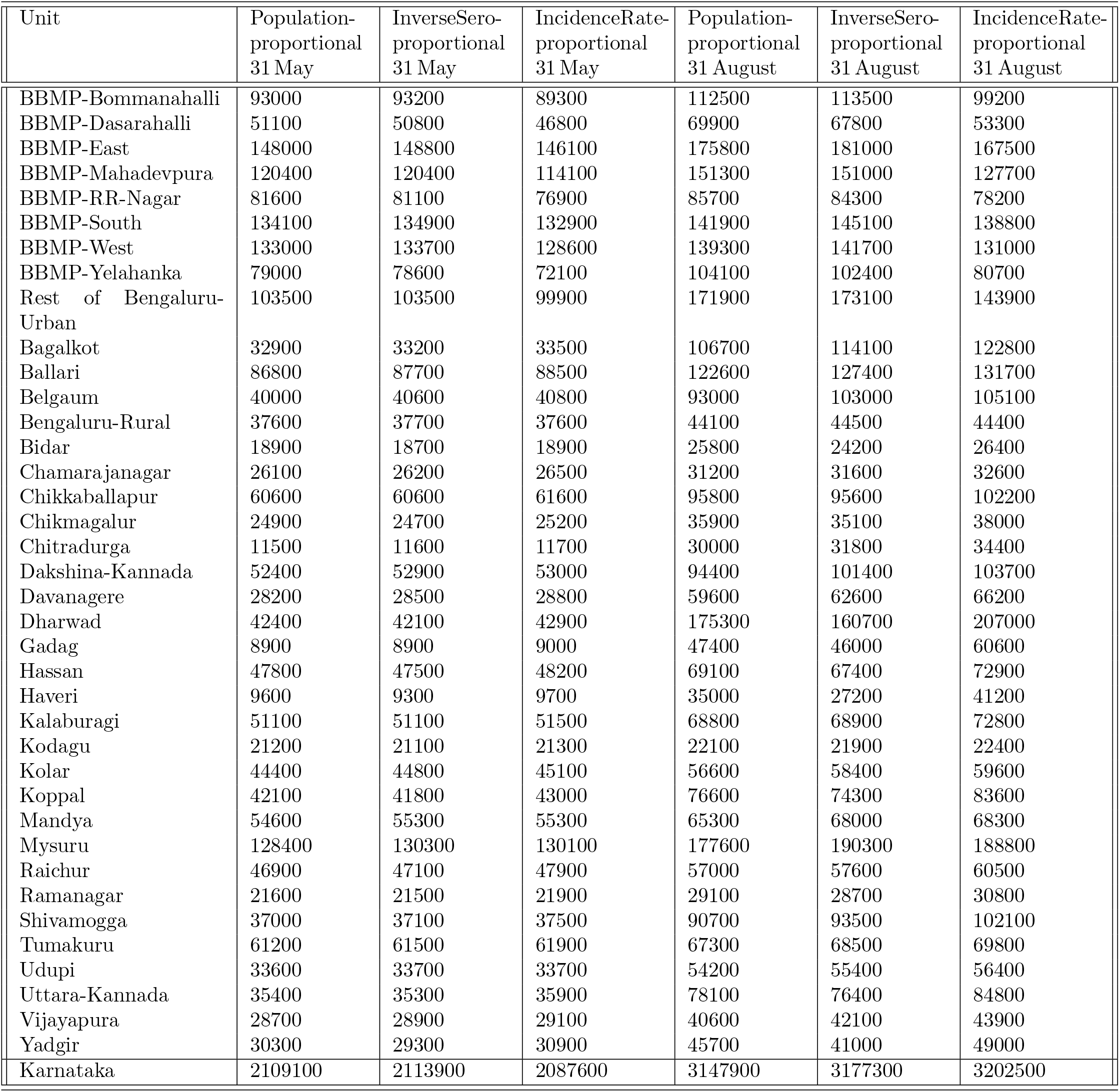
Comparison of unit-wise cumulative reported cases (since 01 November 2020) on 31 May 2021 and 31 August 2021 under No-NPI with a daily budget of 167000 vaccine doses.

| Unit | Population-proportional<br>31 May | InverseSero-proportional<br>31 May | IncidenceRate-proportional<br>31 May | Population-proportional<br>31 August | InverseSero-proportional<br>31 August | IncidenceRate-proportional<br>31 August |
| --- | --- | --- | --- | --- | --- | --- |
| BBMP-Bommanahalli | 93000 | 93200 | 89300 | 112500 | 113500 | 99200 |
| BBMP-Dasarahalli | 51100 | 50800 | 46800 | 69900 | 67800 | 53300 |
| BBMP-East | 148000 | 148800 | 146100 | 175800 | 181000 | 167500 |
| BBMP-Mahadevpura | 120400 | 120400 | 114100 | 151300 | 151000 | 127700 |
| BBMP-RR-Nagar | 81600 | 81100 | 76900 | 85700 | 84300 | 78200 |
| BBMP-South | 134100 | 134900 | 132900 | 141900 | 145100 | 138800 |
| BBMP-West | 133000 | 133700 | 128600 | 139300 | 141700 | 131000 |
| BBMP-Yelahanka | 79000 | 78600 | 72100 | 104100 | 102400 | 80700 |
| Rest of Bengaluru-Urban | 103500 | 103500 | 99900 | 171900 | 173100 | 143900 |
| Bagalkot | 32900 | 33200 | 33500 | 106700 | 114100 | 122800 |
| Ballari | 86800 | 87700 | 88500 | 122600 | 127400 | 131700 |
| Belgaum | 40000 | 40600 | 40800 | 93000 | 103000 | 105100 |
| Bengaluru-Rural | 37600 | 37700 | 37600 | 44100 | 44500 | 44400 |
| Bidar | 18900 | 18700 | 18900 | 25800 | 24200 | 26400 |
| Chamarajanagar | 26100 | 26200 | 26500 | 31200 | 31600 | 32600 |
| Chikkaballapur | 60600 | 60600 | 61600 | 95800 | 95600 | 102200 |
| Chikmagalur | 24900 | 24700 | 25200 | 35900 | 35100 | 38000 |
| Chitradurga | 11500 | 11600 | 11700 | 30000 | 31800 | 34400 |
| Dakshina-Kannada | 52400 | 52900 | 53000 | 94400 | 101400 | 103700 |
| Davanagere | 28200 | 28500 | 28800 | 59600 | 62600 | 66200 |
| Dharwad | 42400 | 42100 | 42900 | 175300 | 160700 | 207000 |
| Gadag | 8900 | 8900 | 9000 | 47400 | 46000 | 60600 |
| Hassan | 47800 | 47500 | 48200 | 69100 | 67400 | 72900 |
| Haveri | 9600 | 9300 | 9700 | 35000 | 27200 | 41200 |
| Kalaburagi | 51100 | 51100 | 51500 | 68800 | 68900 | 72800 |
| Kodagu | 21200 | 21100 | 21300 | 22100 | 21900 | 22400 |
| Kolar | 44400 | 44800 | 45100 | 56600 | 58400 | 59600 |
| Koppal | 42100 | 41800 | 43000 | 76600 | 74300 | 83600 |
| Mandya | 54600 | 55300 | 55300 | 65300 | 68000 | 68300 |
| Mysuru | 128400 | 130300 | 130100 | 177600 | 190300 | 188800 |
| Raichur | 46900 | 47100 | 47900 | 57000 | 57600 | 60500 |
| Ramanagar | 21600 | 21500 | 21900 | 29100 | 28700 | 30800 |
| Shivamogga | 37000 | 37100 | 37500 | 90700 | 93500 | 102100 |
| Tumakuru | 61200 | 61500 | 61900 | 67300 | 68500 | 69800 |
| Udupi | 33600 | 33700 | 33700 | 54200 | 55400 | 56400 |
| Uttara-Kannada | 35400 | 35300 | 35900 | 78100 | 76400 | 84800 |
| Vijayapura | 28700 | 28900 | 29100 | 40600 | 42100 | 43900 |
| Yadgir | 30300 | 29300 | 30900 | 45700 | 41000 | 49000 |
| Karnataka | 2109100 | 2113900 | 2087600 | 3147900 | 3177300 | 3202500 |

**Table 5:** Comparison of unit-wise cumulative reported cases (since 01 November 2020) on 31 May 2021 and 31 August 2021 under 1/3-NPI with a daily budget of 167000 vaccine doses.

| Unit | Population-proportional<br>31 May | InverseSero-proportional<br>31 May | IncidenceRate-proportional<br>31 May | Population-proportional<br>31 August | InverseSero-proportional<br>31 August | IncidenceRate-proportional<br>31 August |
| --- | --- | --- | --- | --- | --- | --- |
| BBMP-Bommanahalli | 79900 | 79900 | 77900 | 90400 | 90900 | 83900 |
| BBMP-Dasarahalli | 42100 | 41900 | 39900 | 50800 | 50000 | 43300 |
| BBMP-East | 129100 | 129500 | 128100 | 143400 | 145600 | 139800 |
| BBMP-Mahadevpura | 102900 | 102800 | 99700 | 117300 | 117300 | 106900 |
| BBMP-RR-Nagar | 76500 | 76100 | 73600 | 79000 | 78300 | 74400 |
| BBMP-South | 125400 | 125900 | 124700 | 129800 | 131200 | 128300 |
| BBMP-West | 125400 | 125800 | 122700 | 129000 | 130100 | 124400 |
| BBMP-Yelahanka | 65700 | 65400 | 62200 | 77900 | 77200 | 66900 |
| Rest of Bengaluru-Urban | 79500 | 79500 | 77900 | 111500 | 112300 | 99200 |
| Bagalkot | 21300 | 21400 | 21500 | 51700 | 56000 | 60700 |
| Ballari | 63600 | 64000 | 64400 | 93400 | 97500 | 100900 |
| Belgaum | 26900 | 27100 | 27200 | 54400 | 61000 | 62400 |
| Bengaluru-Rural | 32500 | 32600 | 32600 | 36300 | 36500 | 36400 |
| Bidar | 16600 | 16500 | 16600 | 18400 | 18000 | 18500 |
| Chamarajanagar | 21000 | 21000 | 21200 | 26000 | 26300 | 27100 |
| Chikkaballapur | 42400 | 42400 | 42900 | 70600 | 70800 | 76200 |
| Chikmagalur | 19100 | 19000 | 19200 | 26000 | 25500 | 27200 |
| Chitradurga | 8500 | 8500 | 8500 | 14800 | 15600 | 16600 |
| Dakshina-Kannada | 40800 | 41000 | 41000 | 56500 | 59000 | 59800 |
| Davanagere | 18900 | 19000 | 19200 | 37600 | 39900 | 42500 |
| Dharwad | 30100 | 30000 | 30300 | 56500 | 53900 | 62600 |
| Gadag | 6500 | 6500 | 6600 | 11000 | 10900 | 12000 |
| Hassan | 37700 | 37600 | 37900 | 49200 | 48400 | 51200 |
| Haveri | 6300 | 6200 | 6400 | 14900 | 12100 | 17700 |
| Kalaburagi | 43400 | 43400 | 43600 | 50000 | 50100 | 51200 |
| Kodagu | 19500 | 19400 | 19500 | 20400 | 20300 | 20600 |
| Kolar | 35400 | 35500 | 35700 | 44600 | 45900 | 46700 |
| Koppal | 28000 | 27800 | 28300 | 52800 | 51400 | 58600 |
| Mandya | 45300 | 45600 | 45700 | 53300 | 55100 | 55300 |
| Mysuru | 100000 | 101000 | 100800 | 132300 | 140300 | 139400 |
| Raichur | 37100 | 37100 | 37600 | 47000 | 47500 | 49900 |
| Ramanagar | 16100 | 16000 | 16300 | 22700 | 22500 | 24100 |
| Shivamogga | 25900 | 25900 | 26100 | 47700 | 49300 | 53200 |
| Tumakuru | 54500 | 54600 | 54900 | 58900 | 59500 | 60300 |
| Udupi | 26500 | 26500 | 26600 | 34700 | 35200 | 35600 |
| Uttara-Kannada | 24000 | 23900 | 24200 | 47400 | 46700 | 52100 |
| Vijayapura | 22500 | 22600 | 22700 | 29400 | 30300 | 31200 |
| Yadgir | 21300 | 20800 | 21600 | 34300 | 30800 | 37200 |
| Karnataka | 1718000 | 1720000 | 1705400 | 2221800 | 2249100 | 2254200 |

**Table 6:** Comparison of unit-wise cumulative reported cases (since 01 November 2020) on 31 May 2021 and 31 August 2021 under 1/2-NPI with a daily budget of 167000 vaccine doses.

| Unit | Population-proportional<br>31 May | InverseSero-proportional<br>31 May | IncidenceRate-proportional<br>31 May | Population-proportional<br>31 August | InverseSero-proportional<br>31 August | IncidenceRate-proportional<br>31 August |
| --- | --- | --- | --- | --- | --- | --- |
| BBMP-Bommanahalli | 73700 | 73800 | 72500 | 79500 | 79700 | 76100 |
| BBMP-Dasarahalli | 38100 | 38000 | 36700 | 42300 | 42000 | 38700 |
| BBMP-East | 120300 | 120500 | 119600 | 128100 | 129100 | 126300 |
| BBMP-Mahadevpura | 95000 | 95000 | 93000 | 102400 | 102400 | 97200 |
| BBMP-RR-Nagar | 73800 | 73600 | 71700 | 75400 | 75000 | 72400 |
| BBMP-South | 120900 | 121300 | 120500 | 123700 | 124500 | 122800 |
| BBMP-West | 121300 | 121600 | 119500 | 123700 | 124300 | 120700 |
| BBMP-Yelahanka | 59800 | 59800 | 57700 | 66000 | 65800 | 60400 |
| Rest of Bengaluru-Urban | 69700 | 69800 | 68900 | 83800 | 84200 | 79000 |
| Bagalkot | 17200 | 17300 | 17300 | 27200 | 28400 | 29700 |
| Ballari | 52600 | 52900 | 53200 | 71200 | 73700 | 75700 |
| Belgaum | 22000 | 22200 | 22200 | 33000 | 35400 | 35900 |
| Bengaluru-Rural | 30100 | 30200 | 30200 | 32300 | 32400 | 32300 |
| Bidar | 15600 | 15600 | 15600 | 16400 | 16300 | 16400 |
| Chamarajanagar | 18100 | 18200 | 18300 | 21800 | 22000 | 22500 |
| Chikkaballapur | 34500 | 34500 | 34700 | 50900 | 51100 | 54200 |
| Chikmagalur | 16500 | 16500 | 16600 | 20100 | 19900 | 20700 |
| Chitradurga | 7400 | 7400 | 7400 | 9500 | 9700 | 9900 |
| Dakshina-Kannada | 36200 | 36300 | 36400 | 42500 | 43300 | 43600 |
| Davanagere | 15400 | 15400 | 15500 | 23600 | 24600 | 25600 |
| Dharwad | 25800 | 25800 | 26000 | 33400 | 32900 | 34700 |
| Gadag | 5700 | 5700 | 5700 | 6900 | 6900 | 7200 |
| Hassan | 33400 | 33300 | 33500 | 39000 | 38700 | 39900 |
| Haveri | 5200 | 5200 | 5300 | 7800 | 7100 | 8400 |
| Kalaburagi | 40200 | 40200 | 40300 | 43300 | 43300 | 43700 |
| Kodagu | 18300 | 18300 | 18400 | 19100 | 19000 | 19200 |
| Kolar | 30900 | 31000 | 31100 | 36600 | 37300 | 37700 |
| Koppal | 22200 | 22200 | 22400 | 35200 | 34600 | 38300 |
| Mandya | 40500 | 40800 | 40800 | 45600 | 46600 | 46700 |
| Mysuru | 87300 | 87800 | 87700 | 104900 | 108700 | 108200 |
| Raichur | 31600 | 31700 | 32000 | 38900 | 39300 | 40800 |
| Ramanagar | 13400 | 13400 | 13500 | 17700 | 17600 | 18600 |
| Shivamogga | 21800 | 21800 | 21900 | 29700 | 30200 | 31300 |
| Tumakuru | 50900 | 51000 | 51200 | 53800 | 54200 | 54600 |
| Udupi | 23700 | 23700 | 23700 | 27200 | 27400 | 27500 |
| Uttara-Kannada | 19700 | 19600 | 19800 | 29400 | 29200 | 31200 |
| Vijayapura | 19700 | 19800 | 19900 | 23300 | 23700 | 24100 |
| Yadgir | 17200 | 16900 | 17400 | 25200 | 23200 | 27000 |
| Karnataka | 1546100 | 1548000 | 1538200 | 1790400 | 1803300 | 1799100 |

**Table 7:** Comparison of unit-wise cumulative reported cases (since 01 November 2020) on 31 May 2021 and 31 August 2021 under 2/3-NPI with a daily budget of 167000 vaccine doses.

| Unit | Population-proportional<br>31 May | InverseSero-proportional<br>31 May | IncidenceRate-proportional<br>31 May | Population-proportional<br>31 August | InverseSero-proportional<br>31 August | IncidenceRate-proportional<br>31 August |
| --- | --- | --- | --- | --- | --- | --- |
| BBMP-Bommanahalli | 68200 | 68200 | 67500 | 70700 | 70700 | 69200 |
| BBMP-Dasarahalli | 34800 | 34700 | 34000 | 36400 | 36200 | 34900 |
| BBMP-East | 112400 | 112500 | 112000 | 115800 | 116100 | 115000 |
| BBMP-Mahadevpura | 88100 | 88100 | 86900 | 91200 | 91100 | 88900 |
| BBMP-RR-Nagar | 71200 | 71000 | 69800 | 72000 | 71800 | 70200 |
| BBMP-South | 116600 | 116800 | 116300 | 118100 | 118500 | 117600 |
| BBMP-West | 117400 | 117600 | 116200 | 118700 | 119000 | 117000 |
| BBMP-Yelahanka | 54900 | 54700 | 53600 | 57300 | 57000 | 54800 |
| Rest of Bengaluru-Urban | 61800 | 61900 | 61400 | 66400 | 66500 | 65100 |
| Bagalkot | 14200 | 14200 | 14200 | 16500 | 16600 | 16800 |
| Ballari | 43300 | 43400 | 43500 | 50300 | 51000 | 51500 |
| Belgaum | 18300 | 18400 | 18400 | 21100 | 21500 | 21600 |
| Bengaluru-Rural | 27900 | 27900 | 27900 | 28800 | 28900 | 28900 |
| Bidar | 14800 | 14800 | 14800 | 15100 | 15100 | 15100 |
| Chamarajanagar | 15400 | 15500 | 15500 | 17100 | 17300 | 17400 |
| Chikkaballapur | 28000 | 28000 | 28100 | 33400 | 33400 | 34200 |
| Chikmagalur | 14400 | 14400 | 14400 | 15700 | 15600 | 15800 |
| Chitradurga | 6600 | 6600 | 6600 | 7100 | 7100 | 7100 |
| Dakshina-Kannada | 32500 | 32600 | 32600 | 34500 | 34700 | 34800 |
| Davanagere | 12600 | 12700 | 12700 | 14800 | 15000 | 15200 |
| Dharwad | 22600 | 22600 | 22600 | 24600 | 24400 | 24700 |
| Gadag | 5100 | 5100 | 5100 | 5400 | 5400 | 5400 |
| Hassan | 29700 | 29700 | 29700 | 31700 | 31600 | 31900 |
| Haveri | 4400 | 4400 | 4400 | 5000 | 4800 | 5000 |
| Kalaburagi | 37400 | 37400 | 37500 | 38600 | 38600 | 38800 |
| Kodagu | 17100 | 17100 | 17100 | 17600 | 17500 | 17600 |
| Kolar | 26900 | 26900 | 27000 | 29200 | 29500 | 29600 |
| Koppal | 17700 | 17600 | 17800 | 21500 | 21300 | 22100 |
| Mandya | 36100 | 36200 | 36200 | 38400 | 38800 | 38800 |
| Mysuru | 76100 | 76400 | 76300 | 82700 | 83800 | 83600 |
| Raichur | 26600 | 26600 | 26700 | 29900 | 30100 | 30600 |
| Ramanagar | 11000 | 11000 | 11100 | 12700 | 12700 | 13000 |
| Shivamogga | 18700 | 18700 | 18700 | 20800 | 20900 | 21000 |
| Tumakuru | 47400 | 47400 | 47500 | 48800 | 49000 | 49200 |
| Udupi | 21300 | 21300 | 21300 | 22500 | 22500 | 22600 |
| Uttara-Kannada | 16400 | 16300 | 16400 | 18900 | 18800 | 19200 |
| Vijayapura | 17400 | 17500 | 17500 | 18700 | 18800 | 18900 |
| Yadgir | 13800 | 13700 | 13800 | 16600 | 16000 | 17000 |
| Karnataka | 1399200 | 1399800 | 1393300 | 1484600 | 1487900 | 1480300 |

### Comparison of the three vaccination policies

Table 5 provides a summary of the cumulative model-projected reported cases on 15 May 2021 and 31 August 2021, respectively, starting from a reference level taken to be zero on 01 November 2020 (which is the reference date)^5^. This is under the three vaccination policies assuming that (i) the daily budget of available vaccine doses is 167000 and (ii) 1/3-NPI is in force starting from 02 May 2021. The (short-term) model-projected cumulative number of reported cases in the state on 31 May 2021, starting from 01 November 2020, is 1.718, 1.720 and 1.705 million under the vaccination policies of the population proportional, inverse seroprevalence proportional and case-incidence rate proportional strategies, respectively. This suggests that all three vaccination strategies are similarly effective in the short term according to the model. Similarly, the model-projected cumulative reported cases on 31 August 2021 are 2.222 million, 2.249 million and 2.254 million under the same three policies, respectively. While the above was for 1/3-NPI, Table 4, Table 6 and Table 7 summarise the model-projected cases under No-NPI, 1/2-NPI and 2/3-NPI, respectively. All of these are within 2% of each other. However, a finer observation indicates that the incidence-rate proportional allocation is more effective in the short term, but the population-proportional allocation is effective in the long-term when viewed at the level of the state^6^.

In the BBMP units, the incidence-rate proportional allocation continues to remain the most effective among the three.

### The benefit of increased daily vaccinations

Table 2 summarises the effect of increased/decreased number of daily vaccinations under various NPIs, assuming that vaccines are allocated in proportion to population. Under No-NPI, the second column in Table 2, we find that the cumulative number of projected cases on 31 August 2021 reduces from 3.27 million to 2.82 million as the number of daily vaccinations increases from 100000 to 400000. Reductions are observed in the other NPIs as well. Going row-wise, for a fixed daily vaccination budget, we find a significant reduction in the number of cumulative cases as NPIs are strengthened, as expected. Going column-wise, an increase of vaccination capacity is most effective when the intensity of NPI is low. Table 3 summarises the reductions in percentage with respect to the reference number of cases under No-NPI and 100000 vaccinations per day.

### Resurgence

Table 8 shows the susceptible population percentage (model-predicted) in each unit. We note that with 100000 vaccination capacity per day, 15*/*38 units have more than 60% susceptible on 31 August 2021. See also Figure 7 which shows that the susceptible fraction increases during August-November 2021 due to antibody waning. Figure 6 shows a future peak between mid-November and mid-December 2021. Figure 11 shows the resurgence in the units. Except for Bidar and Kalaburagi, all show resurgence before December 2021. In our model, Bidar and Kalaburagi will show resurgence since the infection is never wiped out in the simulator. The resurgence in our model can be attributed to residual levels of infection in the unit, which then begins to increase as immunity wanes. Mobility and variants that can escape immunity are not taken into account.

**Table 8:** The unit-wise percentage of susceptible population on 31 August 2021 with varying daily budgets of available vaccine doses under the 1/3-NPI, when vaccines are allocated in proportion to the population of each unit.

| Unit | 100K | 133K | 167K | 200K | 233K | 267K | 300K | 333K | 367K | 400K |
| --- | --- | --- | --- | --- | --- | --- | --- | --- | --- | --- |
| BBMP-Bommanahalli | 59.3 | 55.8 | 52.2 | 48.6 | 45.1 | 41.4 | 37.9 | 34.2 | 30.5 | 27.1 |
| BBMP-Dasarahalli | 61.7 | 58.3 | 54.8 | 51.1 | 47.6 | 44.0 | 40.4 | 36.8 | 33.1 | 29.6 |
| BBMP-East | 60.6 | 57.1 | 53.4 | 49.9 | 46.3 | 42.5 | 38.9 | 35.4 | 31.6 | 28.1 |
| BBMP-Mahadevpura | 60.6 | 57.1 | 53.5 | 49.9 | 46.3 | 42.6 | 39.0 | 35.3 | 31.5 | 28.6 |
| BBMP-RR-Nagar | 55.2 | 51.5 | 47.7 | 44.1 | 40.5 | 36.7 | 32.9 | 29.6 | 27.5 | 26.1 |
| BBMP-South | 59.0 | 55.3 | 51.6 | 47.9 | 44.3 | 40.5 | 36.8 | 33.1 | 29.4 | 26.9 |
| BBMP-West | 56.3 | 52.7 | 49.0 | 45.3 | 41.5 | 37.8 | 34.1 | 30.5 | 28.1 | 26.7 |
| BBMP-Yelahanka | 60.0 | 56.4 | 52.9 | 49.3 | 45.8 | 42.1 | 38.5 | 35.0 | 31.2 | 28.3 |
| Bagalkot | 60.5 | 57.6 | 54.6 | 51.6 | 48.5 | 45.3 | 42.1 | 38.9 | 35.6 | 32.3 |
| Ballari | 47.2 | 44.2 | 41.1 | 37.9 | 34.8 | 31.5 | 28.3 | 25.0 | 22.1 | 20.1 |
| Belgaum | 56.8 | 53.9 | 51.0 | 48.0 | 45.0 | 41.8 | 38.6 | 35.5 | 32.1 | 28.9 |
| Bengaluru-Rural | 57.6 | 54.1 | 50.5 | 47.0 | 43.3 | 39.6 | 36.0 | 32.5 | 28.8 | 25.9 |
| Rest of Bengaluru-Urban | 62.0 | 58.8 | 55.4 | 52.0 | 48.7 | 45.2 | 41.7 | 38.3 | 34.6 | 31.2 |
| Bidar | 74.0 | 70.3 | 66.4 | 62.7 | 58.9 | 55.1 | 51.3 | 47.5 | 43.7 | 39.9 |
| Chamarajanagar | 45.0 | 41.8 | 38.5 | 35.3 | 32.0 | 28.6 | 25.3 | 22.3 | 20.6 | 19.5 |
| Chikkaballapur | 45.4 | 42.5 | 39.5 | 36.5 | 33.4 | 30.3 | 27.2 | 24.3 | 22.0 | 20.4 |
| Chikmagalur | 52.5 | 49.2 | 45.9 | 42.6 | 39.2 | 35.8 | 32.4 | 29.3 | 26.9 | 25.1 |
| Chitradurga | 64.8 | 61.6 | 58.3 | 55.0 | 51.7 | 48.3 | 44.9 | 41.5 | 37.9 | 34.5 |
| Dakshina-Kannada | 64.4 | 61.0 | 57.5 | 54.0 | 50.5 | 46.9 | 43.4 | 39.8 | 36.2 | 32.9 |
| Davanagere | 53.7 | 50.9 | 48.0 | 45.1 | 42.1 | 39.0 | 35.9 | 32.8 | 29.5 | 26.3 |
| Dharwad | 73.8 | 70.3 | 66.7 | 63.1 | 59.6 | 55.9 | 52.3 | 48.7 | 44.9 | 41.3 |
| Gadag | 78.2 | 74.5 | 70.6 | 66.9 | 63.2 | 59.3 | 55.5 | 51.8 | 47.9 | 44.1 |
| Hassan | 56.7 | 53.4 | 49.9 | 46.6 | 43.1 | 39.6 | 36.1 | 32.6 | 29.5 | 27.2 |
| Haveri | 66.3 | 63.3 | 60.1 | 57.0 | 53.8 | 50.5 | 47.2 | 43.9 | 40.4 | 37.1 |
| Kalaburagi | 67.7 | 64.1 | 60.4 | 56.7 | 53.1 | 49.4 | 45.7 | 42.0 | 38.2 | 34.6 |
| Kodagu | 46.4 | 43.1 | 39.4 | 35.9 | 32.4 | 29.2 | 26.6 | 24.8 | 23.3 | 22.2 |
| Kolar | 51.1 | 47.9 | 44.5 | 41.3 | 37.9 | 34.5 | 31.1 | 27.7 | 24.4 | 22.0 |
| Koppal | 48.2 | 45.5 | 42.5 | 39.7 | 36.7 | 33.5 | 30.5 | 27.4 | 24.3 | 21.9 |
| Mandya | 50.4 | 47.0 | 43.5 | 40.0 | 36.6 | 33.1 | 29.6 | 26.5 | 24.1 | 22.3 |
| Mysuru | 50.9 | 47.7 | 44.3 | 41.0 | 37.7 | 34.1 | 31.0 | 28.4 | 26.3 | 24.5 |
| Raichur | 43.2 | 40.0 | 36.7 | 33.5 | 30.2 | 26.8 | 24.1 | 22.0 | 20.3 | 19.3 |
| Ramanagar | 44.5 | 41.4 | 38.2 | 35.1 | 31.9 | 28.6 | 25.5 | 23.0 | 21.2 | 19.5 |
| Shivamogga | 61.6 | 58.6 | 55.3 | 52.1 | 48.9 | 45.5 | 42.1 | 38.8 | 35.3 | 32.1 |
| Tumakuru | 53.3 | 49.8 | 46.3 | 42.7 | 39.1 | 35.5 | 31.9 | 28.3 | 25.0 | 23.7 |
| Udupi | 61.9 | 58.5 | 54.9 | 51.4 | 47.9 | 44.2 | 40.7 | 37.1 | 33.9 | 31.3 |
| Uttara-Kannada | 53.4 | 50.6 | 47.6 | 44.6 | 41.6 | 38.4 | 35.3 | 32.1 | 29.0 | 26.7 |
| Vijayapura | 57.1 | 53.8 | 50.5 | 47.1 | 43.7 | 40.2 | 36.8 | 33.3 | 29.8 | 26.5 |
| Yadgir | 46.8 | 44.0 | 40.9 | 37.9 | 34.9 | 31.7 | 28.6 | 25.5 | 22.1 | 19.5 |
| Karnataka | 57.2 | 53.9 | 50.5 | 47.2 | 43.9 | 40.4 | 37.0 | 33.7 | 30.6 | 27.9 |

**Figure 7:**
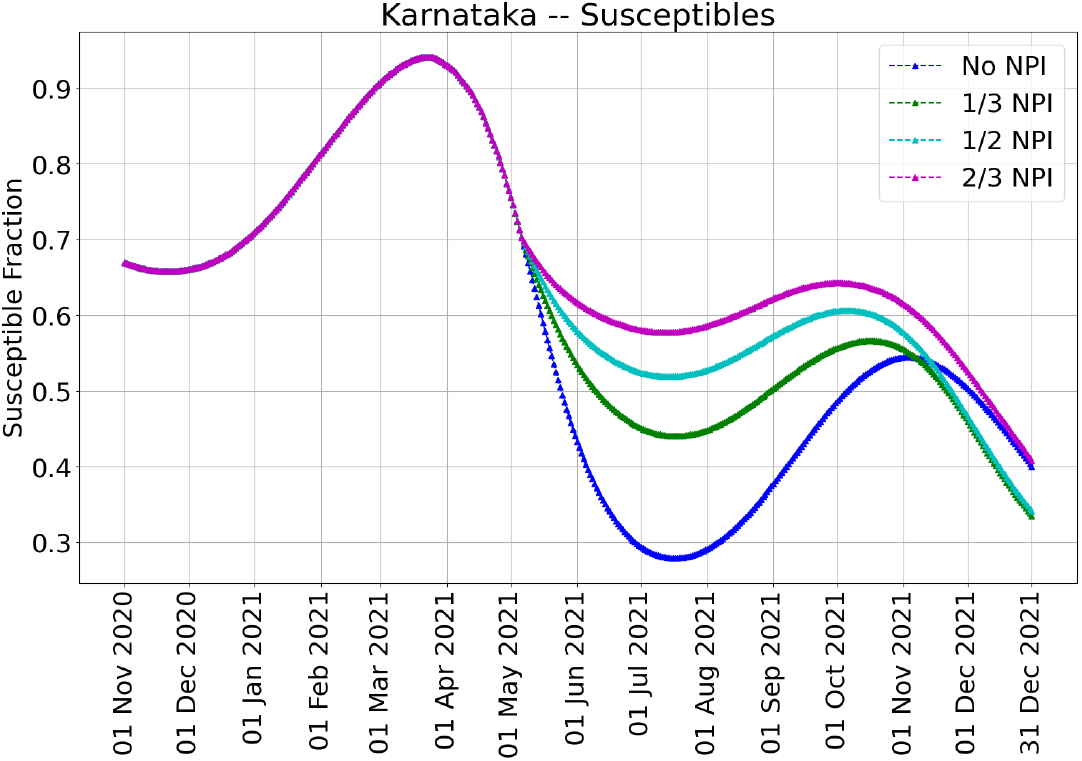
The evolution of susceptible fraction in the state of Karnataka over time. These plots are for the vaccination policy that allocates 167000 vaccine doses per day proportional to each unit’s population. Observe that the susceptible fraction increase during the period August-November 2021 due to antibody waning.

## 4. Discussion and Limitations

We first discuss the implications of our results. We then discuss limitations of our study arising from model errors, uncertainty in the factors driving the resurgence in Karnataka, policy changes in response to the local circumstances, and vaccine supply constraints.

### 4.1 Discussion of the results

The significant second surge has highlighted widespread susceptibility in early 2021. This could be due to waning immunity, or limited spread of the first wave, or novel variants, or a combination thereof. An expedited, effective, and equitable vaccine campaign remains the only feasible pathway to controlling COVID-19 in India. Given supply constraints and India’s population size, tools for designing efficient vaccination campaigns are essential. In this paper, we developed a model (using serosurvey data and on-the-ground datasets) to study vaccination allocation strategies and the interplay of vaccination capacity and NPIs in achieving sufficient immunity levels. The main messages of this work are the following:

- *Heterogeneity across units under various NPI-scenarios:* Our model suggests that there is heterogeneity in the disease spread across units, e.g. under No-NPI the units would have peaked at different times. The statewide uniform NPIs that are in place currently restrict mobility and thus limit the transmission of the virus within/across units. Our model predicts that all units would see a downward trend in reported cases after 15 May 2021. Daily reported cases have indeed come down in most units. Further, compared to No-NPI, on 31 August 2021, the model projects that 1/3-NPI may reduce the (cumulative reported) cases by roughly 926,000. Under 2/3-NPI and 167000 daily vaccinations, the percentage of susceptible populations in the units varies from 36.7% (Raichur) and 70.6% (Gadag), indicating significant heterogeneity across the units (see Table 8).
- *Effectiveness of vaccination strategies:* As for the three vaccination policies, our model indicates that they are roughly equal in the short-term, but in the longer term, statewide allocation in proportion to population leads to fewer cases. This conclusion is robust at the state level across various NPI scenarios. If we focus on units with high incidence in April 2021, e.g. Bengaluru’s BBMP units, these units benefit more from an allocation in proportion to the case-incidence rate. However, all vaccination allocation strategies are within a 2% margin of each other. Given the robustness of the results, population-based allocation strategies are likely to be as successful as any other strategy that may be harder to implement.
- *The interplay of vaccination and NPI on healthcare infrastructure capacity:* With 167000 available vaccine doses per day, we find that the number of cumulative cases on 31 August 2021 reduces from *±* 3.14 million under No-NPI to 1.48 million under 2/3-NPI. Table 3 and Figure 8 quantify the percentage reduction in the number of cases with respect to the reference 100000 vaccine doses/day and No-NPI setting. We have thus identified the key interplay between the intensity of the NPIs and the daily vaccination capacity. Further Table 9 provides the mean ± StdDev of active cases in Karnataka under various NPIs and vaccination allocation strategies during the months of June, July and August 2021. These can be used either at the unit level or at the state level to design a desirable public health response (choice of NPI, vaccination strategy, capacity). For example, if a mean of more than 60,000 active cases in a month would imply that the healthcare infrastructure in Karnataka is operating at full capacity, *then an appropriate public health response starting May 2021 could be to vaccinate at a daily rate of 167000 and choose a social interaction between* 1*/*3*-NPI and* 1*/*2*-NPI. This will help to minimise deaths and manage severity of illness occurring during the surge*.
- *Resurgence:* Our model predicts a resurgence in cases in the form of a third wave during the months of November-December 2021. This prediction is a conservative one since it is based on only seroreversion and mobility being restricted to within units. In reality, the peak could be advanced or delayed due to stochasticity and mobility and could be more prominent due to more transmissible variants.

**Figure 8:**
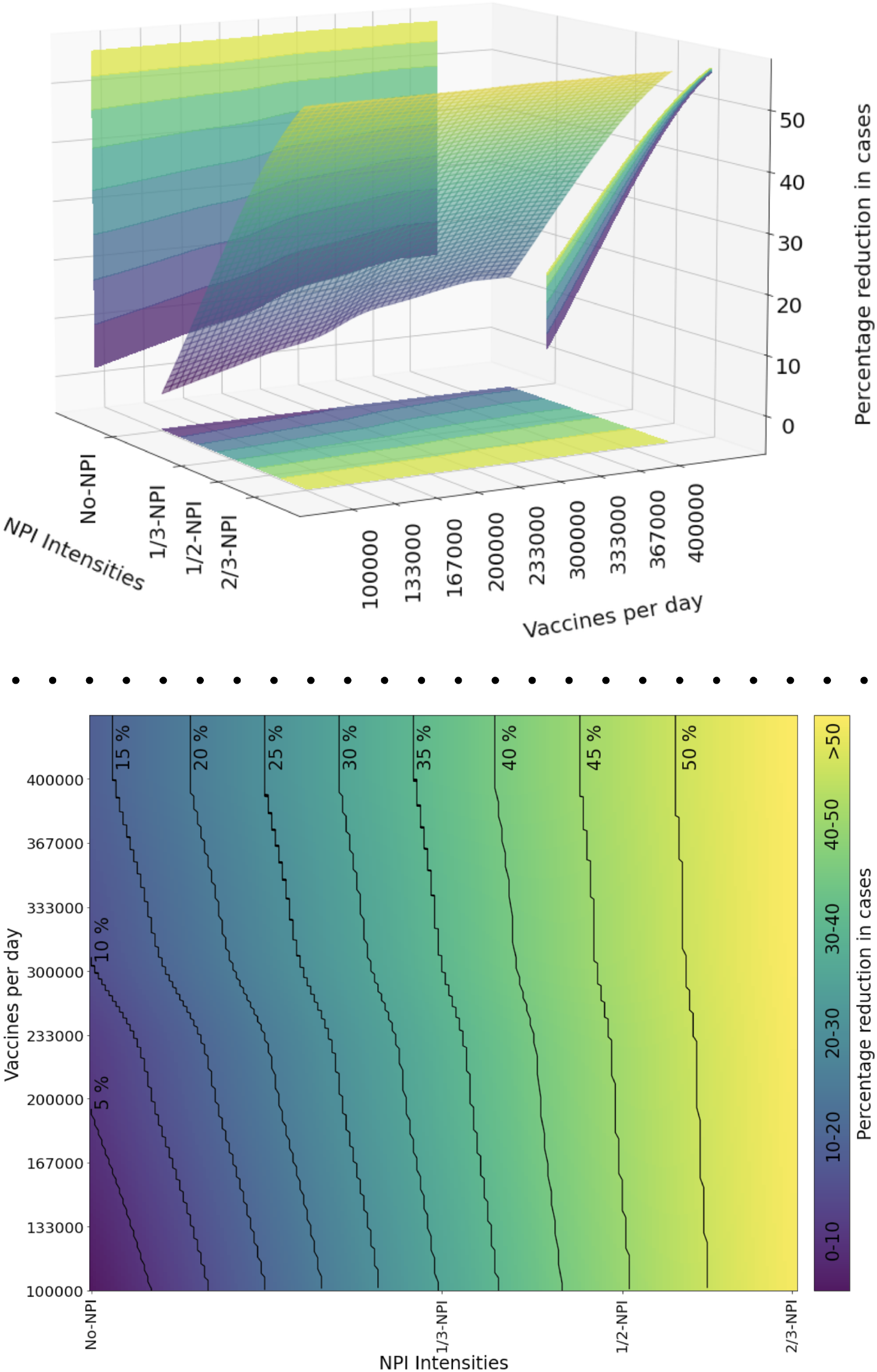
Surface plot (top figure) and contour plot (bottom figure) summarising the reduction in the number of model-projected cases on 31 August 2021 for varying daily vaccination budgets and NPI intensities. The surface plot and the contours are based on linear interpolation from the data points in Table 3.

**Table 9:**
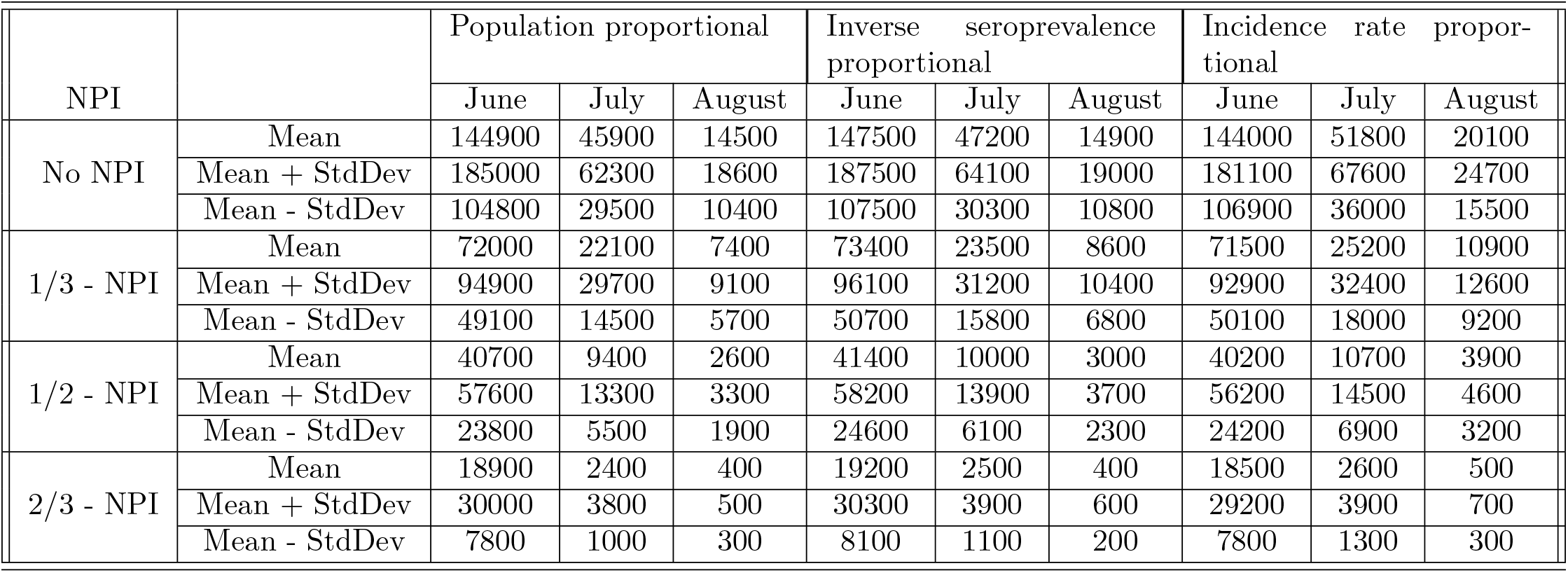
Projected mean StdDev of active cases in Karnataka under various NPIs and vaccination allocation strategies during the months of June, July and August 2021 when the daily vaccination budget is 167000.

| NPI |  | Population proportional |  |  | Inverse seroprevalence proportional |  |  | Incidence rate proportional |  |  |
| --- | --- | --- | --- | --- | --- | --- | --- | --- | --- | --- |
|  |  | June | July | August | June | July | August | June | July | August |
| No NPI | Mean | 144900 | 45900 | 14500 | 147500 | 47200 | 14900 | 144000 | 51800 | 20100 |
|  | Mean + StdDev | 185000 | 62300 | 18600 | 187500 | 64100 | 19000 | 181100 | 67600 | 24700 |
|  | Mean - StdDev | 104800 | 29500 | 10400 | 107500 | 30300 | 10800 | 106900 | 36000 | 15500 |
| 1/3 - NPI | Mean | 72000 | 22100 | 7400 | 73400 | 23500 | 8600 | 71500 | 25200 | 10900 |
|  | Mean + StdDev | 94900 | 29700 | 9100 | 96100 | 31200 | 10400 | 92900 | 32400 | 12600 |
|  | Mean - StdDev | 49100 | 14500 | 5700 | 50700 | 15800 | 6800 | 50100 | 18000 | 9200 |
| 1/2 - NPI | Mean | 40700 | 9400 | 2600 | 41400 | 10000 | 3000 | 40200 | 10700 | 3900 |
|  | Mean + StdDev | 57600 | 13300 | 3300 | 58200 | 13900 | 3700 | 56200 | 14500 | 4600 |
|  | Mean - StdDev | 23800 | 5500 | 1900 | 24600 | 6100 | 2300 | 24200 | 6900 | 3200 |
| 2/3 - NPI | Mean | 18900 | 2400 | 400 | 19200 | 2500 | 400 | 18500 | 2600 | 500 |
|  | Mean + StdDev | 30000 | 3800 | 500 | 30300 | 3900 | 600 | 29200 | 3900 | 700 |
|  | Mean - StdDev | 7800 | 1000 | 300 | 8100 | 1100 | 200 | 7800 | 1300 | 300 |

**Figure 9:**
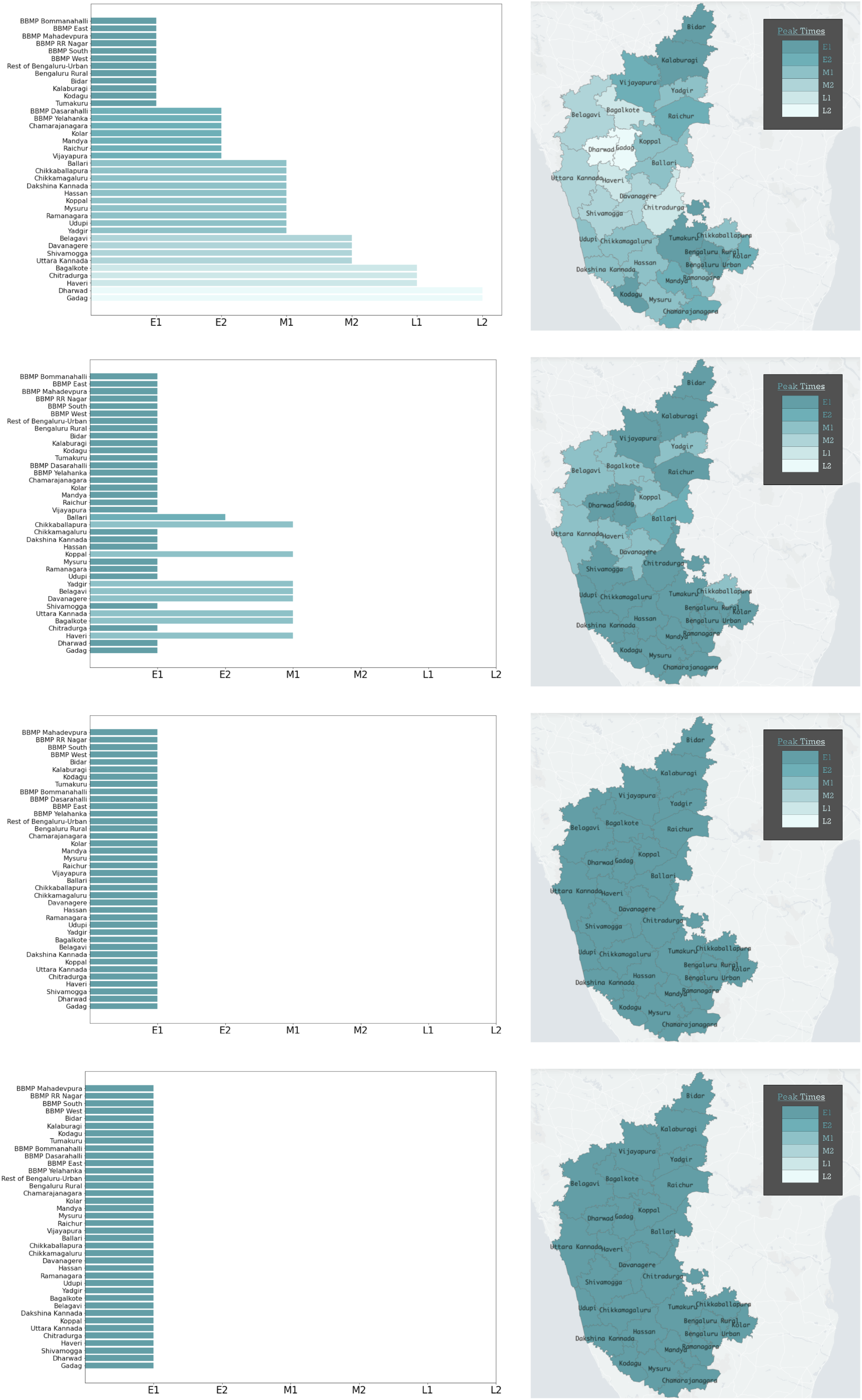
*Left subfigure*: The time of occurrence of the peak number of reported cases for each unit under No-NPI, 1/3-NPI, 1/2-NPI and 2/3-NPI, respectively. E, M, L stand for early, mid, late peaks, and the ordering is: E1 *<* E2 *<* M1 *<* M2 *<* L1 *<* L2. *Right subfigure*: Geo-spatial representations of the two scenarios. Spatial heterogeneity manifests in the No-NPI setting. In the 1/2- and 2/3-NPI settings, the cases start to recede from mid-May 2021. Note that the NPI intensities are held constant until 31 December 2021 in our simulations.

Our model makes use of several real-time data sources – population data, age-distribution, age-stratified contact rates, disease progression data, confirmed case trajectories across units, seroprevalence data, time series of tests conducted, vaccination time-series, and efficacy of vaccines. Our compartmental ODE model used patches that where made up of age and unit stratifications. We modelled mobility across subunits to be independent of age. A future extension could be to use age-stratified mobility data.

There are many models in the Indian context, e.g., [3] with four compartments and [1] with five compartments that are explicitly modelling asymptomatic spread, [40] with nine compartments and explicit modelling of age-stratification and serology-based cases-to-infections ratio factor, and the PDE model in [25]. Our SEIRV spatio-temporal model goes beyond these and includes mobility and seroreversion.

One could also consider agent-based models as in [2], but given the complexity of such a model for Bengaluru alone (12.4 million population), scaling it up six times to the level of Karnataka will involve significant (more-than-linear) complexity increase. Parsimonious SEIRV-based district-level age-stratified ODE models incorporating mobility and seroreversion do provide a good tradeoff point between complexity and the ability to gather insights for public health policy.

Recognising that we already have several parameters in our model – five phases of contact rates and one contact rate per unit in each phase – we used only one constant contact rate per unit and scalings thereof (NPI intensities) for the forward projections. The per-unit contact rates are estimated based on the trajectory of the case during 08 April – 01 May 2021. Further, we quantified uncertainty by doing an estimation during 08–22 April 2021 and by doing a validation from 23 April 2021 – 11 May 2021. Statewide root mean squared estimation error was 9% and included the estimation errors of 40-50% for the units Chikkaballapur, Kodagu, Kolar, Vijaypura, and Raichur. The large errors in these units seem to be due a change in the trend of the number of reported cases which could not be captured by the model.

In terms of the vaccination policy, one could also employ simulation optimisation approaches to derive, algorithmically, vaccination schedules that minimise various epidemiologically relevant objectives under budget constraints as done in [32,46]. Such models use simulation in the loop and can incorporate a forecast horizon at which the vaccination policies are evaluated. When combined with data on vaccine acceptance [31], population profile of co-morbidities [14] and logistical challenges in setting up mass/mobile vaccination sites, one can obtain a realistic spatial distribution and allocation strategy that is executable, effective and equitable.

### 4.2 Limitations

Our first limitation is on the calibration of parameters – we have many contact rate parameters, one for each unit and for each of the five different time horizons. Tuning all of them to match the ground truth may lead to overfitting and loss of predictive power but still provides sufficient ground for scenario-based comparisons.

The selection of time points for contact rate calibration is based on observed regime changes for the trajectory of the case in each unit (see Appendix 4.2). A more systematic approach to arrive at these change points automatically can make the projections scalable to other states and the districts of India. Further, the change in transmissibility, if attributable to an increase in the prevalence of the variant strains (e.g.,or B.1.617.2), can be modelled using estimates of transmissibility advantage [15].

Despite NPIs, infections may continue to rise for some time. The impact of such interventions will manifest only after some time if there are reporting delays. Our model used a delay of 1 week, assuming a two-day sample collection delay and 5-day test-reporting delay. More data is needed to quantify these delays. Another reason for the delayed impact of NPI could be household spread due to ineffective home isolation. Until the home infections naturally go down, these infections may continue to rise. Such effects are best modelled in agent-based simulators.

The exact cause of the resurgence is yet unclear. It could be due to antibody decay, parameters for which are not yet fully understood, and associated reinfections. It could also be due to reinfections arising from variants having modified spike protein configurations offering immune escape or a combination of these reasons. As the recent studies using the Manaus, Brazil outbreak show [9,39], disentangling these factors may be complex without additional data. Our approach to handling these factors is to move recovered individuals into the susceptible pool based on best available information on waning immunity and to increase the contact rates to capture some of the effects of novel variants; these explain the resurgence to some extent. A more rigorous but also a more complex approach would be to bring in more compartments that represent disease states associated with variants and to model variants and their transmissibility.

Our study is predicated on a constant daily vaccination capacity (which varied from 100000 - 400000). However, there are significant vaccine supply shortages and day-to-day variations. This shortage had become particularly acute starting 01 May 2021 when those above 18 years of age became eligible for vaccination. Further, some of the vaccination capacity is set aside for the prescribed second dose, an aspect which we have not modelled in this work. Finally, the current vaccination campaign does not utilise the serostatus of individuals as in [8], and so some proportion of the capacity is used up for those already immune. In our implementation, for simplicity, vaccinations are focused on the susceptibles and optimistically moves 66% of the daily vaccinated number into the vaccinated compartment after 14 days. Further study is required to account for vaccine wastage and immunisation of those already infected.

## Data Availability

Data used is from publicly available data sources.

https://des.kar.nic.in/docs/ProjectedPopulation2012-2021.pdf

## Appendix

### More on the calibration and the piece-wise constant contact rate periods

Our entire model is a spatio-temporal age-stratified SEIRV compartmental model with mobility and antibody waning. To determine the periods when the contact rates can be held constant, let us first turn to a simplified model to make some insightful observations.

#### Decay and growth phases

Let us consider a simplified setting where we ignore mobility, age-stratified interaction, vaccination and antibody waning. Focus on fractions of the population in each compartment and write *s*_*j*_(*t*) = *S*_*j*_(*t*)*/N*_*j*_ for the susceptible fraction in unit *j* at time *t*; similarly define *e*_*j*_(*t*), *i*_*j*_(*t*), *r*_*j*_(*t*) in unit *j* at time *t*. Then the difference equations in (1)-(4) separate for each unit *j*. Let us suppress the unit index *j* in the notation for this section. The difference equations constitute the Euler discretisation for the ordinary differential equation system:

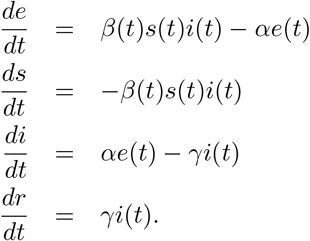

Assuming *β*(*t*) and *s*(*t*) are constant^7^, setting 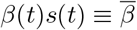, we have the simpler linear system

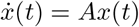

where

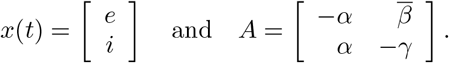

It is easy to verify that *A* is diagonalisable for generic values of *α, β, γ*, and that its two eigenvalues are

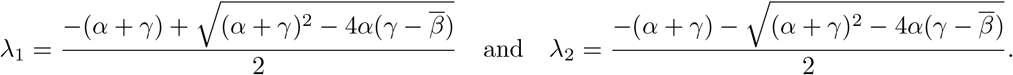

Then *x*(*t*) = *e*^*At*^*x*(0) and hence *i*(*t*) (and also *e*(*t*)) will grow or will decay exponentially as 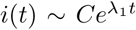, where *λ*_1_ is the larger of the two eigenvalues. This suggests that *λ*_1_ may be estimated as the slope using linear regression on the trajectory log(*i*(*t*)) during the phase when 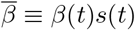 *s*(*t*) is constant. Using standard methods in least squares regression, a 95%-confidence interval for the estimate of *λ*_1_ can be obtained using Student’s *t*-distribution, in this simplified model.

From the plot in Figure 10, we see that the 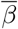 is roughly constant during November 2020 to February 2021, and then from mid-March 2021. These motivate the calibration periods used in this work.

**Figure 10:**
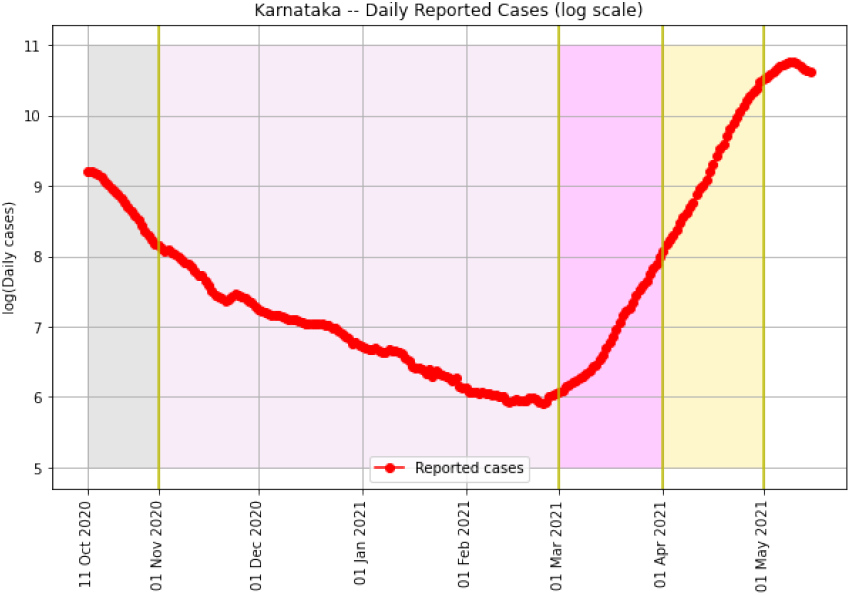
Log plot of the daily infections (averaged over seven days to avoid weekly periodicity issues). The different phases are colour coded. During October 2020, we initialise the simulator. During 01 November 2020 – 28 February 2021, we capture the receding trend. As seen above, both are well-captured by straight lines. During 01 March 2021 – 31 March 2021, we calibrate to re-seed the infections across the units and handle stochasticity. From 01 April 2021 - 01 May 2021, we capture the resurgence and the *λ*1 associated with it. All projections are from 02 May 2021 onward.

The quantity 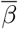 can be recovered from the eigenvalue as:

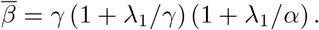

Observe that the reproduction number for this oversimplified model is 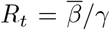, which is given by *R*_*t*_ = (1 + *λ*_1_*/γ*) (1 + *λ*_1_*/α*). Clearly *R*_*t*_ *>* 1 if *λ*_1_ *>* 0 and *R*_*t*_ *<* 1 if *λ*_1_ *<* 0, as expected.

These observations suggest a least-squares regression to estimate *λ*_1_ and then an inversion using the *R*_*t*_ formula to obtain 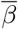. Further, a linear approximation can be used to obtain confidence intervals for 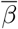 from that of *λ*_1_, in this simplified model.

The actual implementation is a nonlinear least-squares regression that directly searches for the *β*. We take the loss function to be minimised as the squared error objective 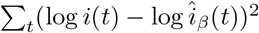 where *î*_*β*_(*t*) is the model-predicted infections at time *t* for contact rate *β*, and the sum is taken over the period for which *β*(·) is assumed constant (called a phase). Note that *î*_*β*_() is a *β*-dependent solution to the difference equations (1)-(4) and respects the actual number of susceptible individuals at each time instant. We implement an iterative gradient descent algorithm to arrive at a good *β* for each unit. Further, since (1)-(4) is a coupled system of patches (units with age-stratification), in each phase, for each iteration, we update the *β*_*j*_ in a round-robin fashion across the units.

#### Calibration before resurgence

Between 01–31 March 2021, we noted in our simulations that while there were residual infections in the Bengaluru units, there was a need to seed infections in some other units to match the timing of the resurgence in these units. The resurgences in these units were likely due to seeding by mobility from other places where the infection was still prevalent. So we model a cross-unit mobility Θ(*t*) = (1 *ε*)*I* + *EJ* for the period of March 2021 with *ε* = 1*/*100. Once seeded, we conservatively model only local transmission, and so we remove the cross-unit mobility from 01 April 2021 onward.

There is also significant stochasticity during 01–31 March 2021. Mean-field models like ours need to be enhanced to handle this stochasticity during such phases. To compensate for the stochasticity, we divide the period into two phases, where we estimate the contact rates. However, the estimation/calibration is only to put the system in a realistic initial state (how many in each of the SEIRV compartments) to learn the best-fit contact rates during the exponential growth phase starting 01 April 2021. Some units see this exponential phase from 15 March 2021 itself, so splitting the month of March into two phases allows some units to fit better the early upsurge (e.g., BBMP units). In summary, the two calibrations during this period is largely to seed infections through mobility, handle stochasticity, and put the system in a more realistic initial state prior to the exponential growth phase starting 01 April 2021.

#### Contact rates

**Table 10:** Calibrated contact rates for each unit. The time periods *T*_1_, …, *T*_5_ are 11–31 October 2020, 01 November 2020 – 28 February 2021, 01–15 March 2021, 16 March – 07 April 2021, 08 April onward, respectively.

| Unit | $T_1$ | $T_2$ | $T_3$ | $T_4$ | $T_5$ |
| --- | --- | --- | --- | --- | --- |
| BBMP-Bommanahalli | 0.018 | 0.025 | 0.074 | 0.04 | 0.047 |
| BBMP-Dasarahalli | 0.016 | 0.022 | 0.056 | 0.038 | 0.042 |
| BBMP-East | 0.041 | 0.022 | 0.06 | 0.043 | 0.047 |
| BBMP-Mahadevpura | 0.013 | 0.021 | 0.062 | 0.042 | 0.043 |
| BBMP-RR-Nagar | 0.019 | 0.023 | 0.092 | 0.043 | 0.062 |
| BBMP-South | 0.033 | 0.026 | 0.084 | 0.048 | 0.058 |
| BBMP-West | 0.032 | 0.029 | 0.08 | 0.047 | 0.062 |
| BBMP-Yelahanka | 0.019 | 0.022 | 0.057 | 0.039 | 0.043 |
| Bagalkot | 0.006 | 0.017 | 0.018 | 0.008 | 0.043 |
| Ballari | 0.01 | 0.025 | 0.057 | 0.015 | 0.056 |
| Belgaum | 0.013 | 0.025 | 0.032 | 0.012 | 0.046 |
| Bengaluru-Rural | 0.019 | 0.025 | 0.061 | 0.046 | 0.049 |
| Rest of Bengaluru-Urban | 0.018 | 0.024 | 0.045 | 0.03 | 0.042 |
| Bidar | 0.019 | 0.025 | 0.045 | 0.045 | 0.029 |
| Chamarajanagar | 0.02 | 0.022 | 0.033 | 0.018 | 0.064 |
| Chikkaballapur | 0.014 | 0.021 | 0.022 | 0.011 | 0.056 |
| Chikmagalur | 0.014 | 0.024 | 0.031 | 0.031 | 0.048 |
| Chitradurga | 0.021 | 0.024 | 0.039 | 0.017 | 0.04 |
| Dakshina-Kannada | 0.014 | 0.025 | 0.054 | 0.029 | 0.039 |
| Davanagere | 0.014 | 0.028 | 0.019 | 0.013 | 0.048 |
| Dharwad | 0.01 | 0.02 | 0.021 | 0.012 | 0.035 |
| Gadag | 0.007 | 0.016 | 0.011 | 0.005 | 0.031 |
| Hassan | 0.017 | 0.023 | 0.047 | 0.035 | 0.045 |
| Haveri | 0.01 | 0.019 | 0.01 | 0.009 | 0.041 |
| Kalaburagi | 0.008 | 0.026 | 0.06 | 0.042 | 0.037 |
| Kodagu | 0.021 | 0.024 | 0.045 | 0.025 | 0.078 |
| Kolar | 0.017 | 0.021 | 0.045 | 0.026 | 0.055 |
| Koppal | 0.015 | 0.016 | 0.007 | 0.01 | 0.052 |
| Mandya | 0.023 | 0.021 | 0.036 | 0.036 | 0.055 |
| Mysuru | 0.017 | 0.024 | 0.047 | 0.026 | 0.05 |
| Raichur | 0.01 | 0.022 | 0.017 | 0.017 | 0.063 |
| Ramanagar | 0.011 | 0.02 | 0.031 | 0.016 | 0.058 |
| Shivamogga | 0.009 | 0.021 | 0.021 | 0.019 | 0.041 |
| Tumakuru | 0.017 | 0.024 | 0.08 | 0.034 | 0.059 |
| Udupi | 0.011 | 0.025 | 0.07 | 0.034 | 0.039 |
| Uttara-Kannada | 0.012 | 0.021 | 0.027 | 0.013 | 0.047 |
| Vijayapura | 0.025 | 0.026 | 0.051 | 0.033 | 0.046 |
| Yadgir | 0.007 | 0.022 | 0.032 | 0.011 | 0.057 |

##### Seroprevalence from the model

**Table 11:** Seroprevalence from the model and the data. Once the second round serosurvey data is available (both active infection and antibody prevalence estimates), the last two columns could be used for comparison and calibration. Number are in percentages.

| Unit | Active Infection %<br>on 18 February 2021<br>(Model) | Antibody(IgG)<br>Prevalence % on<br>18 February 2021<br>(Model) | Antibody (IgG) Preva-<br>lence % (Round-1) |
| --- | --- | --- | --- |
| BBMP-Bommanahalli | 0.039 | 12.359 | 12.5 |
| BBMP-Dasarahalli | 0.045 | 7.987 | 13.8 |
| BBMP-East | 0.068 | 9.906 | 15.5 |
| BBMP-Mahadevpura | 0.044 | 7.176 | 7 |
| BBMP-RR-Nagar | 0.048 | 12.143 | 9.1 |
| BBMP-South | 0.059 | 14.208 | 20.9 |
| BBMP-West | 0.072 | 15.53 | 25.1 |
| BBMP-Yelahanka | 0.05 | 8.404 | 13.9 |
| Bagalkot | 0.004 | 3.987 | 4.1 |
| Ballari | 0.01 | 10.858 | 22.1 |
| Belgaum | 0.04 | 7.84 | 23.7 |
| Bengaluru-Rural | 0.029 | 13.002 | 15.2 |
| Rest of Bengaluru-Urban | 0.046 | 10.825 | 15 |
| Bidar | 0.07 | 4.672 | 18 |
| Chamarajanagar | 0.029 | 8.953 | 15.8 |
| Chikkaballapur | 0.045 | 6.708 | 6.4 |
| Chikmagalur | 0.02 | 12.011 | 12 |
| Chitradurga | 0.044 | 12.621 | 10.2 |
| Dakshina-Kannada | 0.046 | 10.419 | 14.5 |
| Davanagere | 0.019 | 13.527 | 16.4 |
| Dharwad | 0.017 | 2.673 | 7.1 |
| Gadag | 0.001 | 2.101 | 6.3 |
| Hassan | 0.025 | 12.012 | 8.2 |
| Haveri | 0.005 | 8.062 | 14.8 |
| Kalaburagi | 0.053 | 8.975 | 17.1 |
| Kodagu | 0.072 | 12.311 | 12 |
| Kolar | 0.034 | 8.17 | 10.1 |
| Koppal | 0.002 | 4.655 | 19.6 |
| Mandya | 0.014 | 9.631 | 18.5 |
| Mysuru | 0.05 | 9.665 | 18.8 |
| Raichur | 0.007 | 8.097 | 22.8 |
| Ramanagar | 0.006 | 8.735 | 13.9 |
| Shivamogga | 0.023 | 7.514 | 7.7 |
| Tumakuru | 0.045 | 12.424 | 6.8 |
| Udupi | 0.018 | 11.31 | 16.2 |
| Uttara-Kannada | 0.031 | 7.077 | 8.1 |
| Vijayapura | 0.019 | 13.731 | 23.9 |
| Yadgir | 0.01 | 8.551 | 15.4 |

##### Time series of daily and cumulative new infections

**Figure 11:**
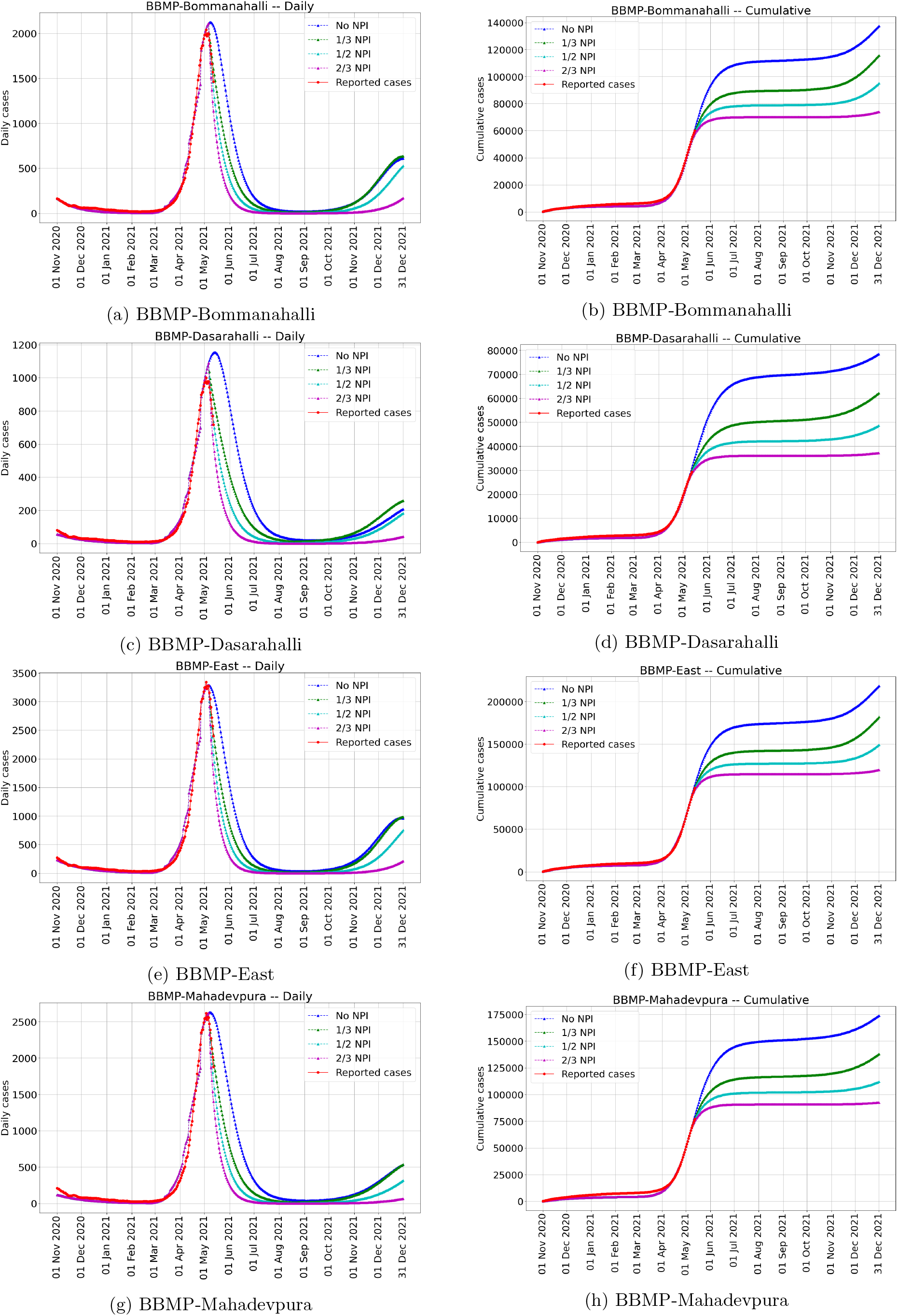
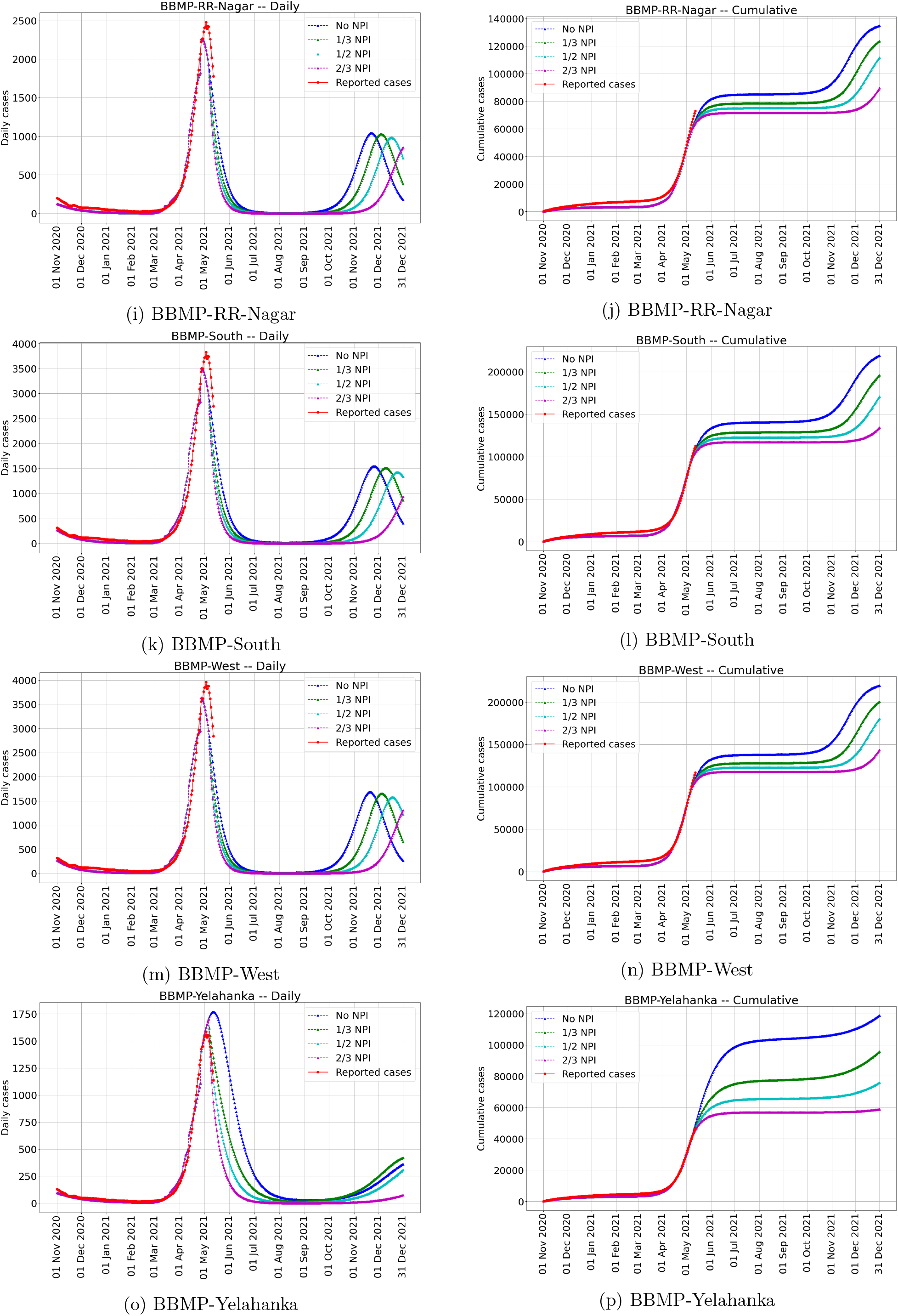
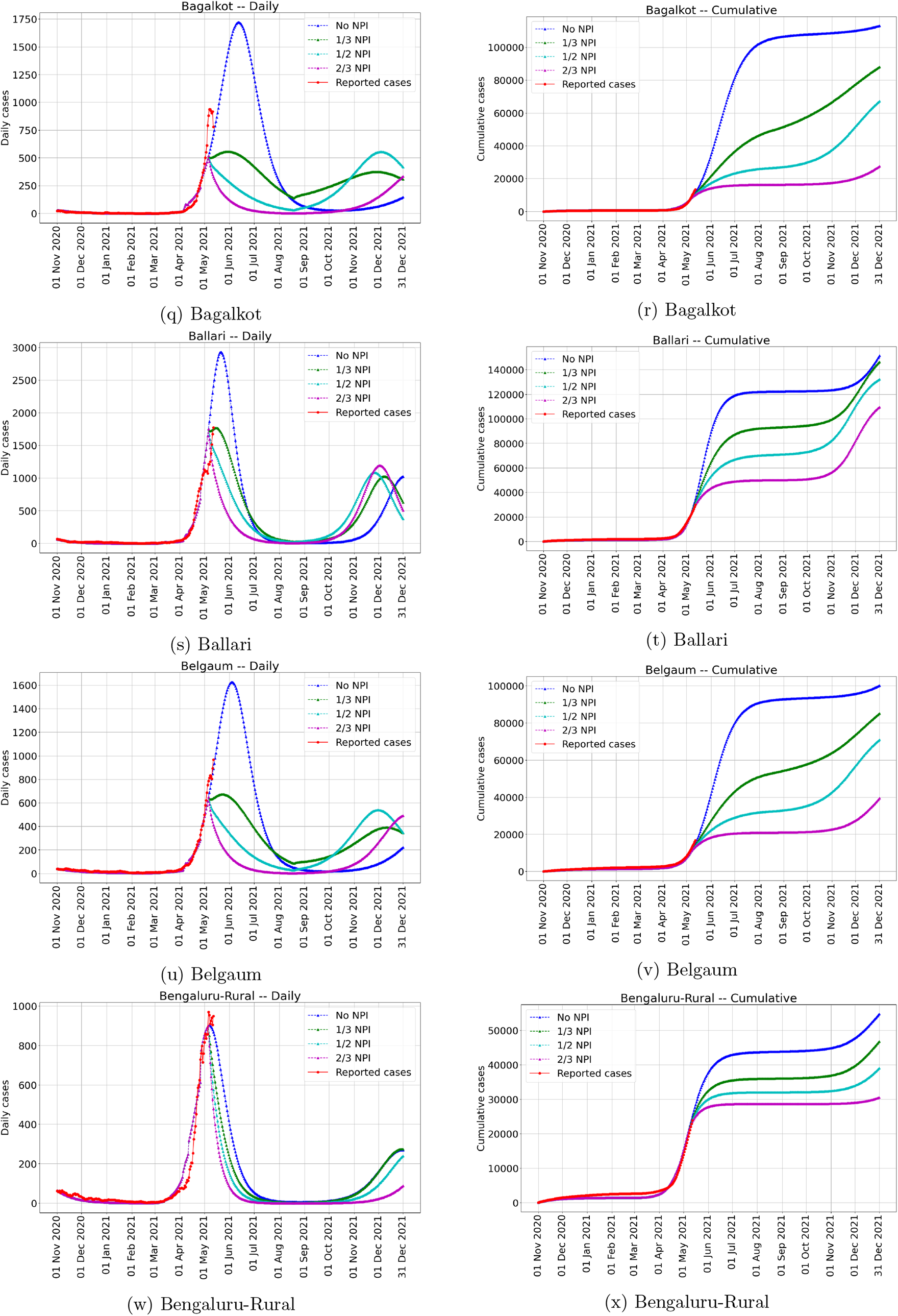
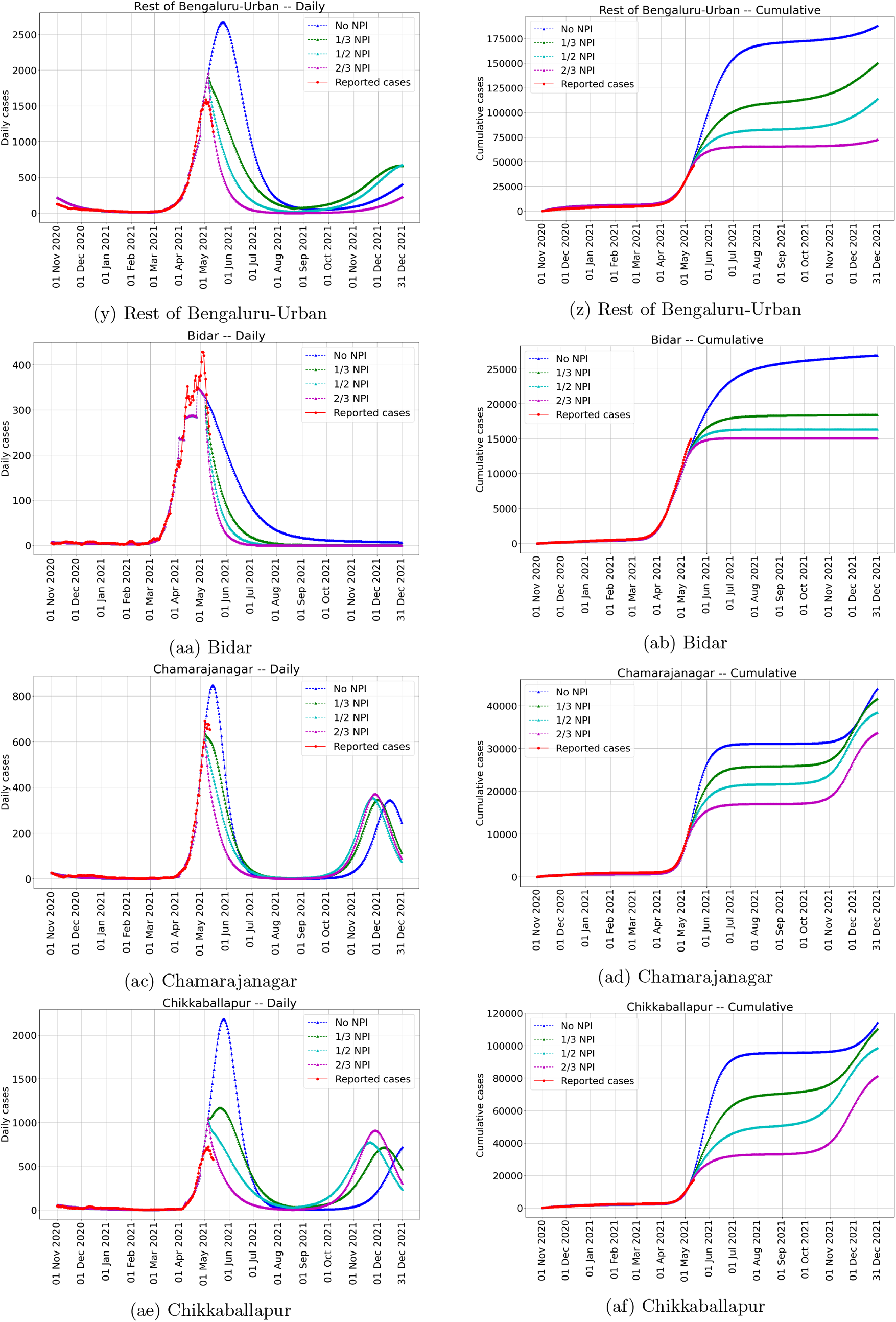
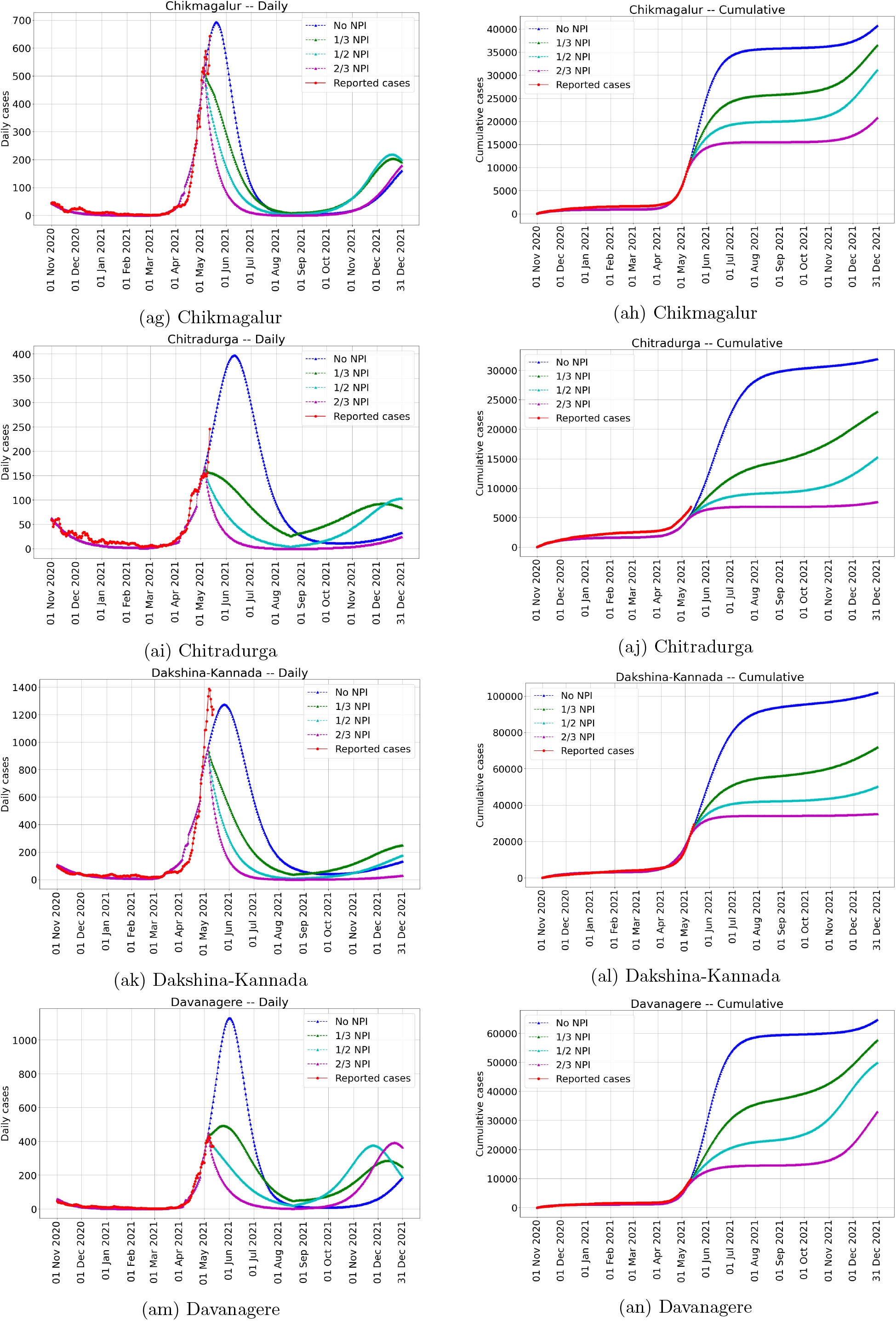
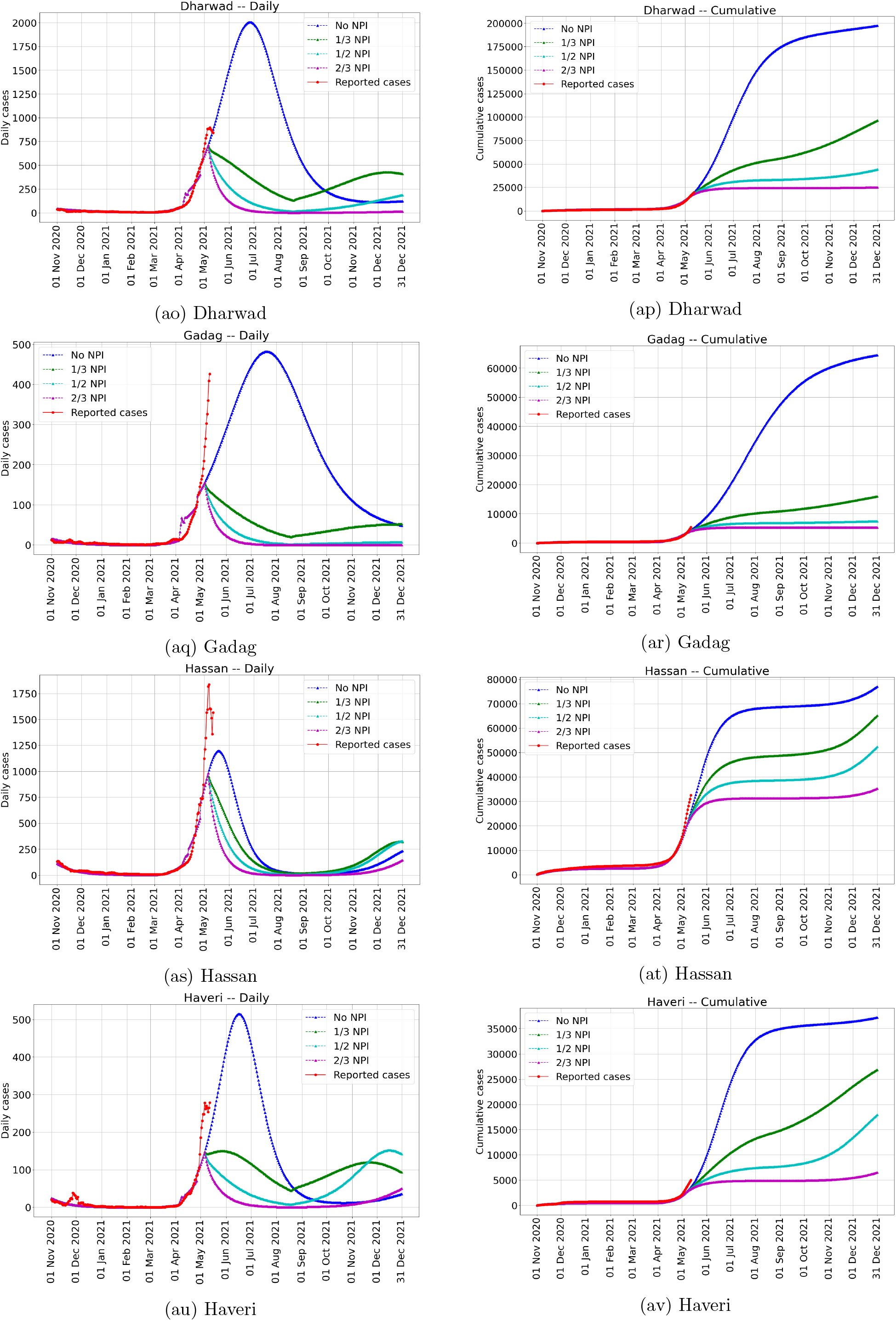
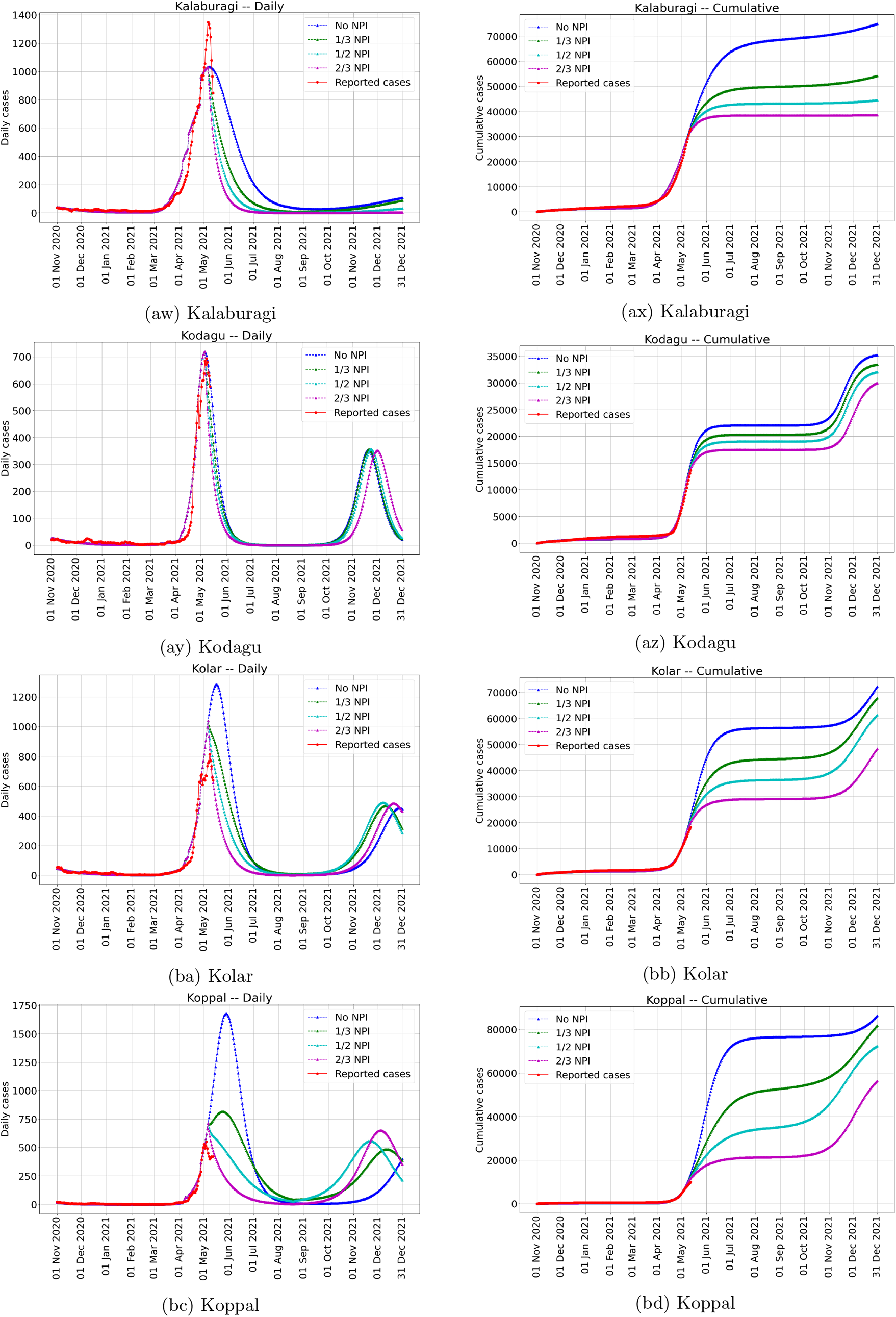
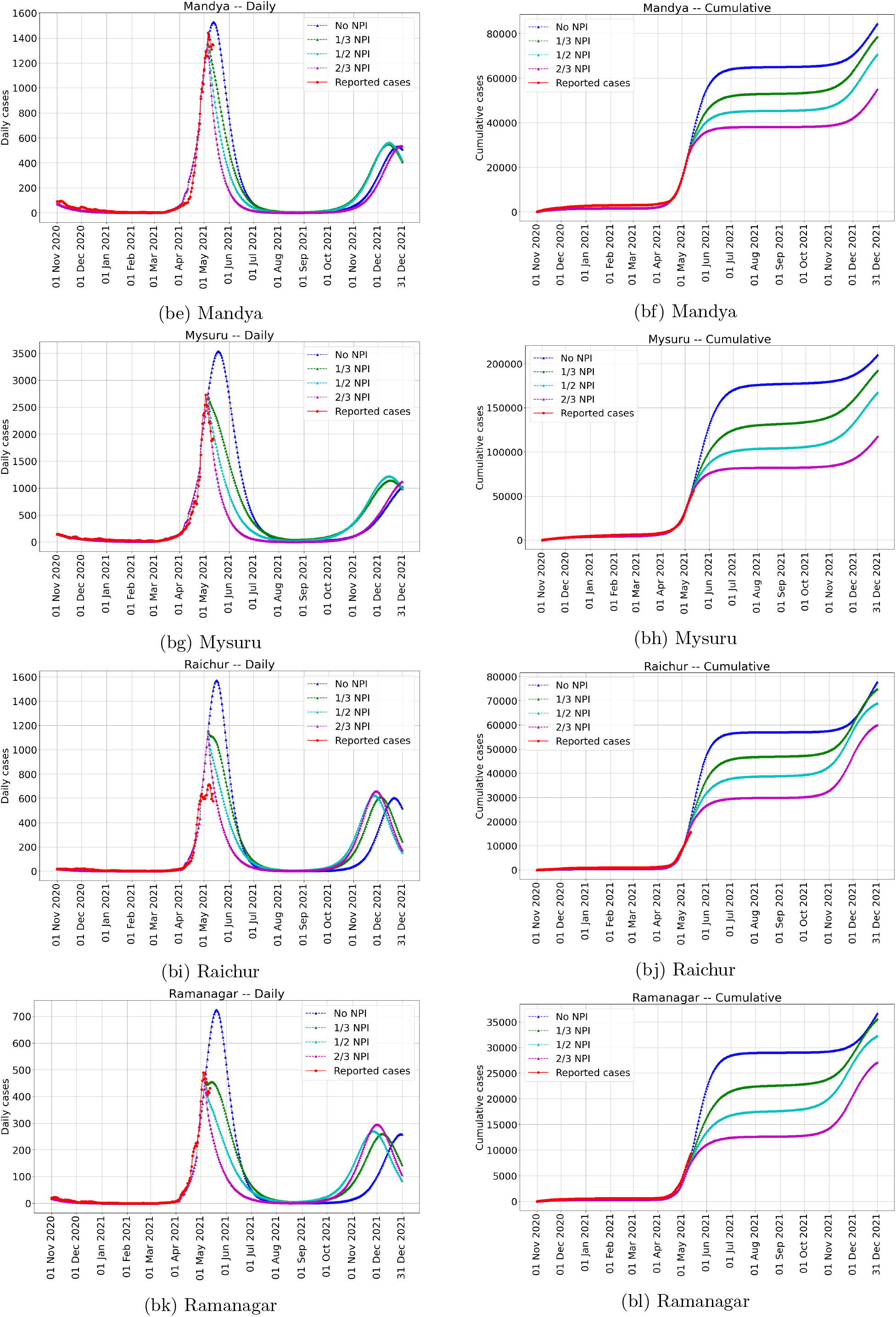
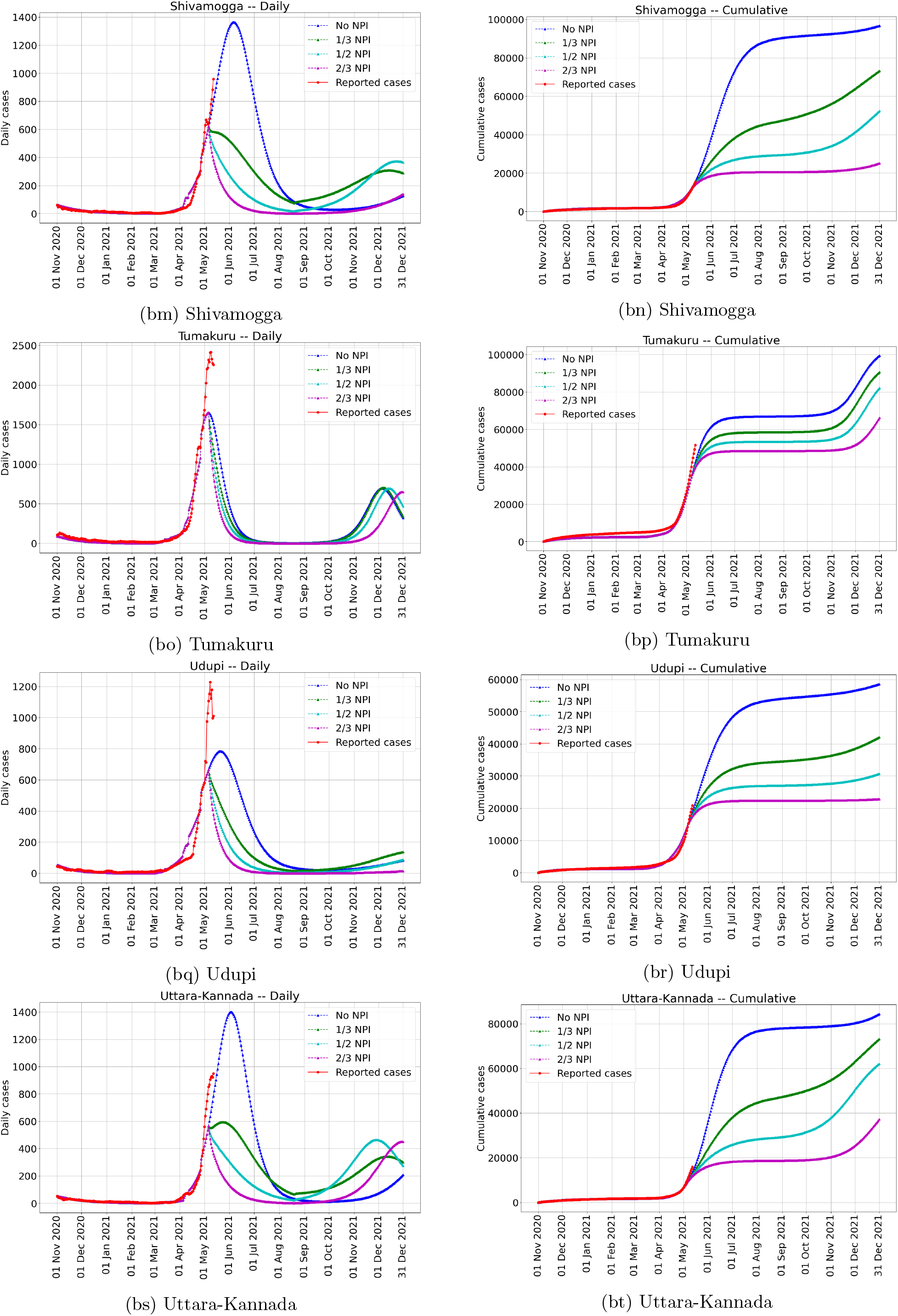
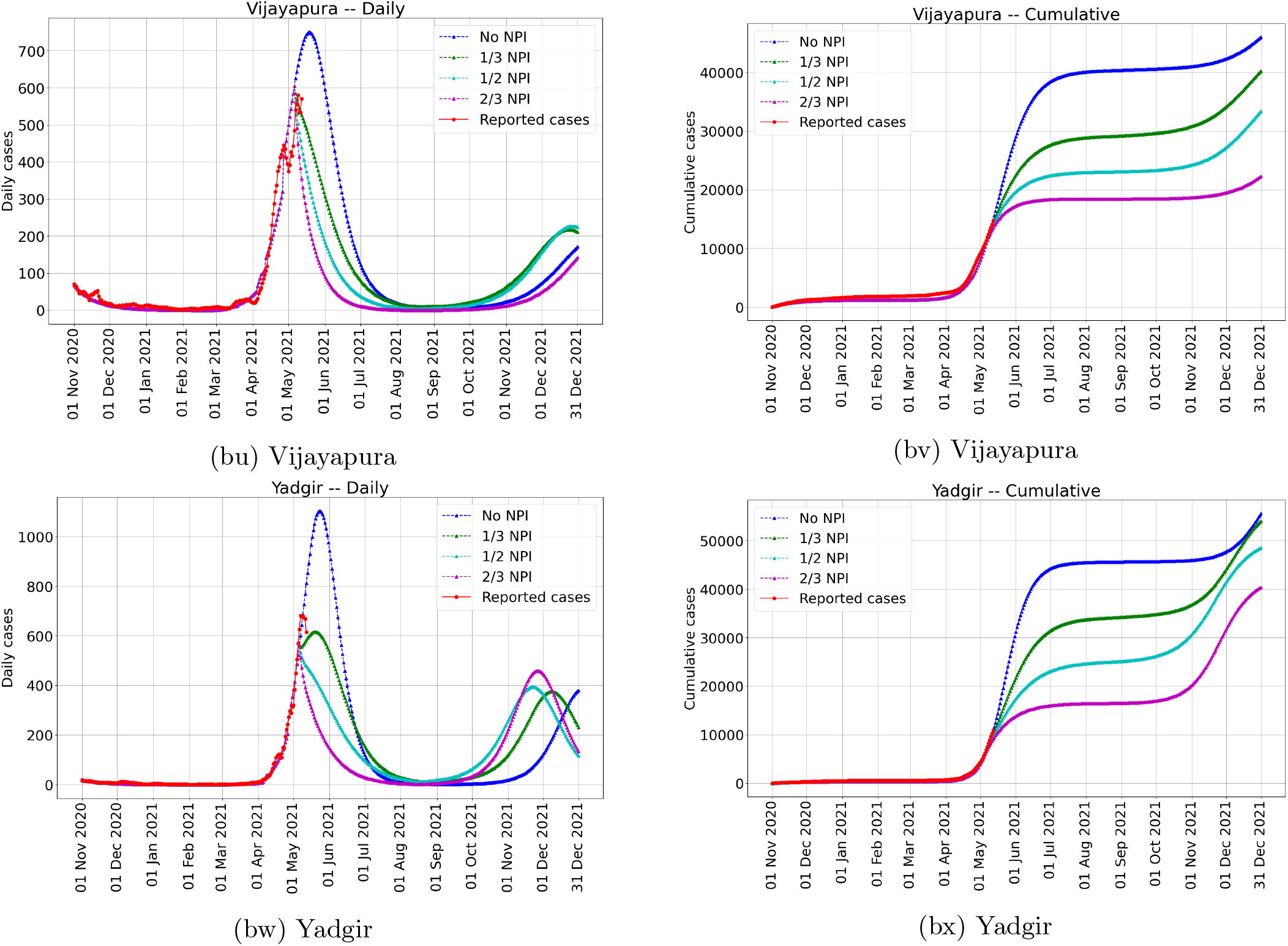
Daily predicted number of cases in each unit until 31 December 2021 for various NPI intensities. Vaccination capacity is 167000 per day allocated in proportion to population in the units.

##### Susceptible fraction

**Figure 12:**
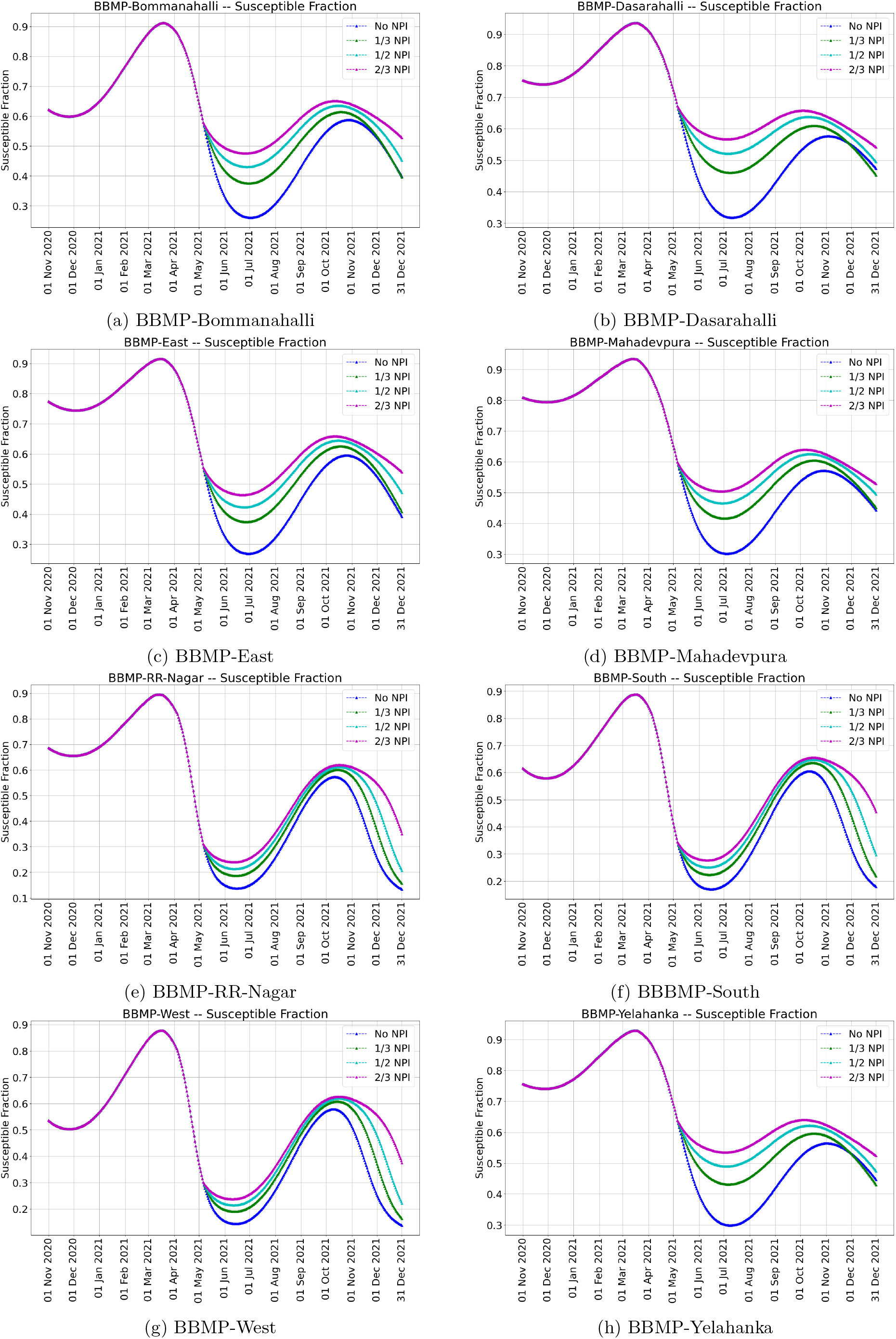
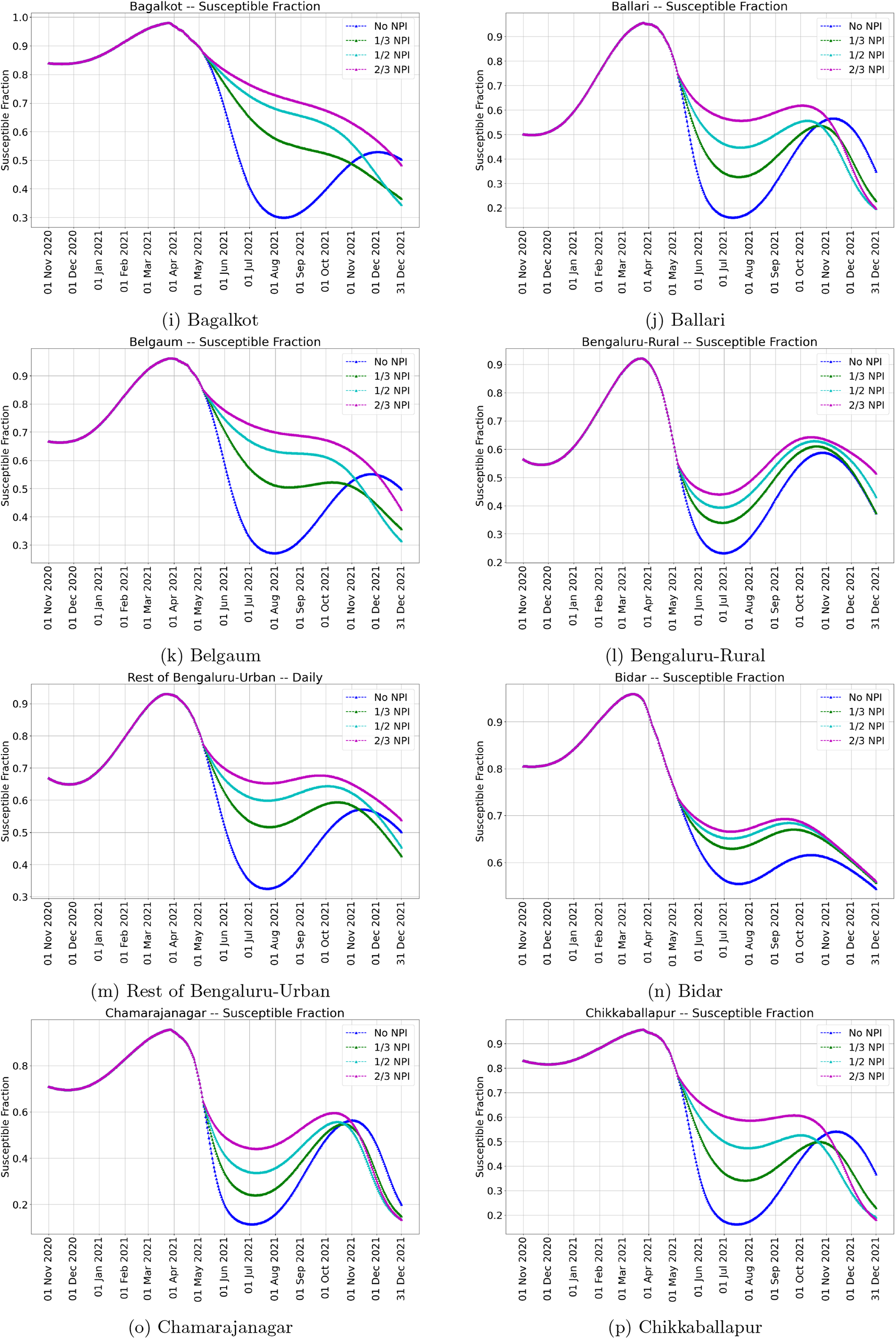
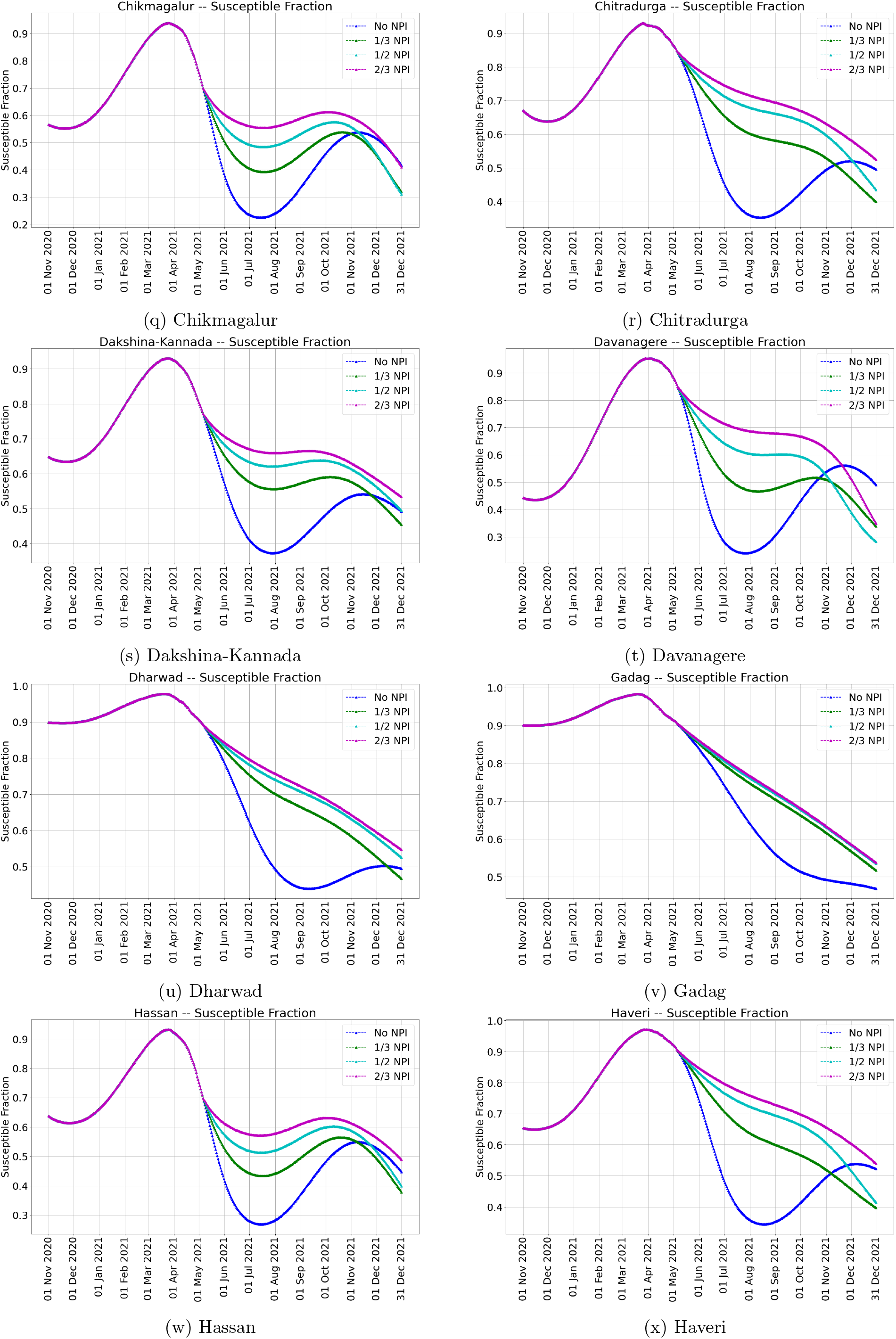
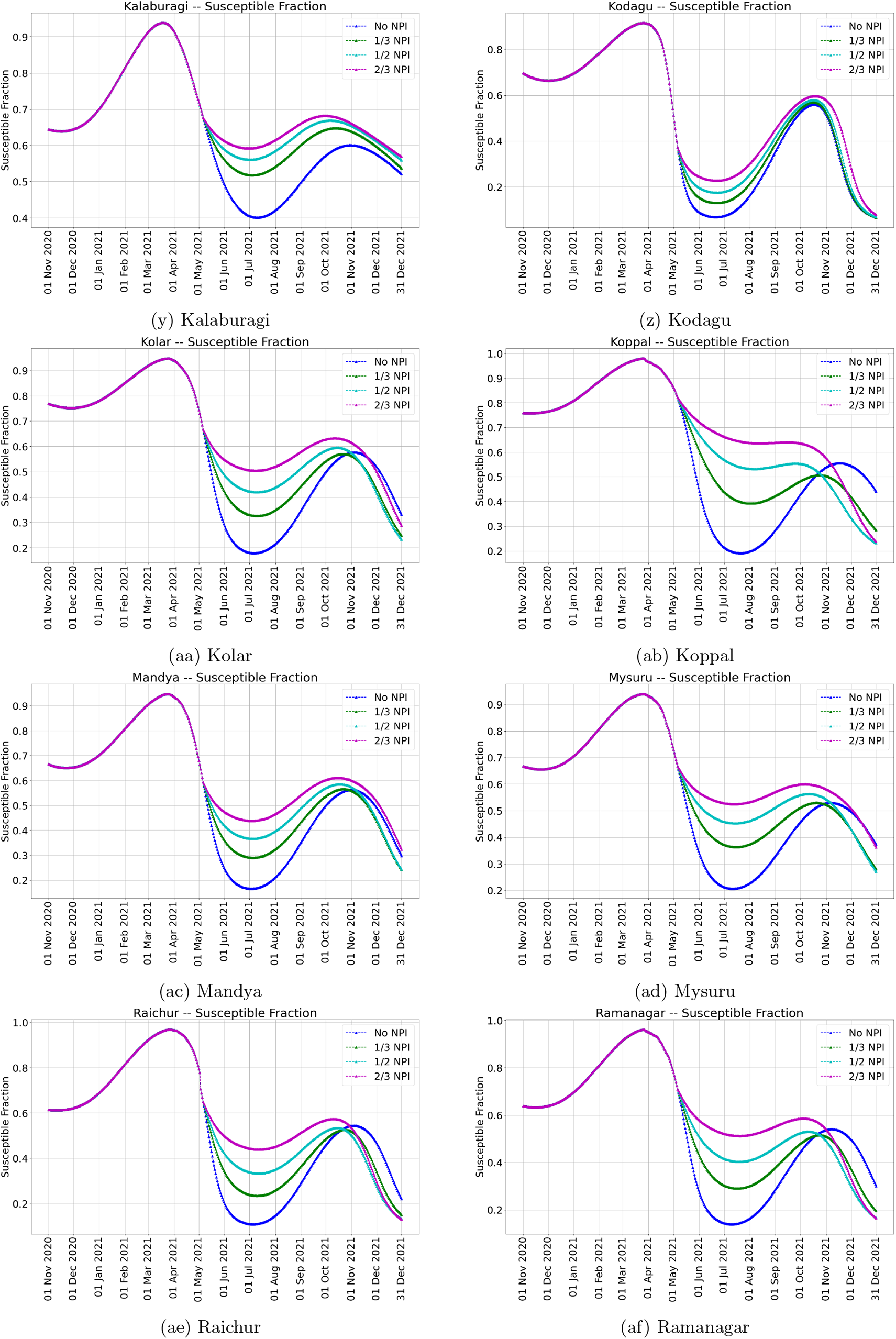
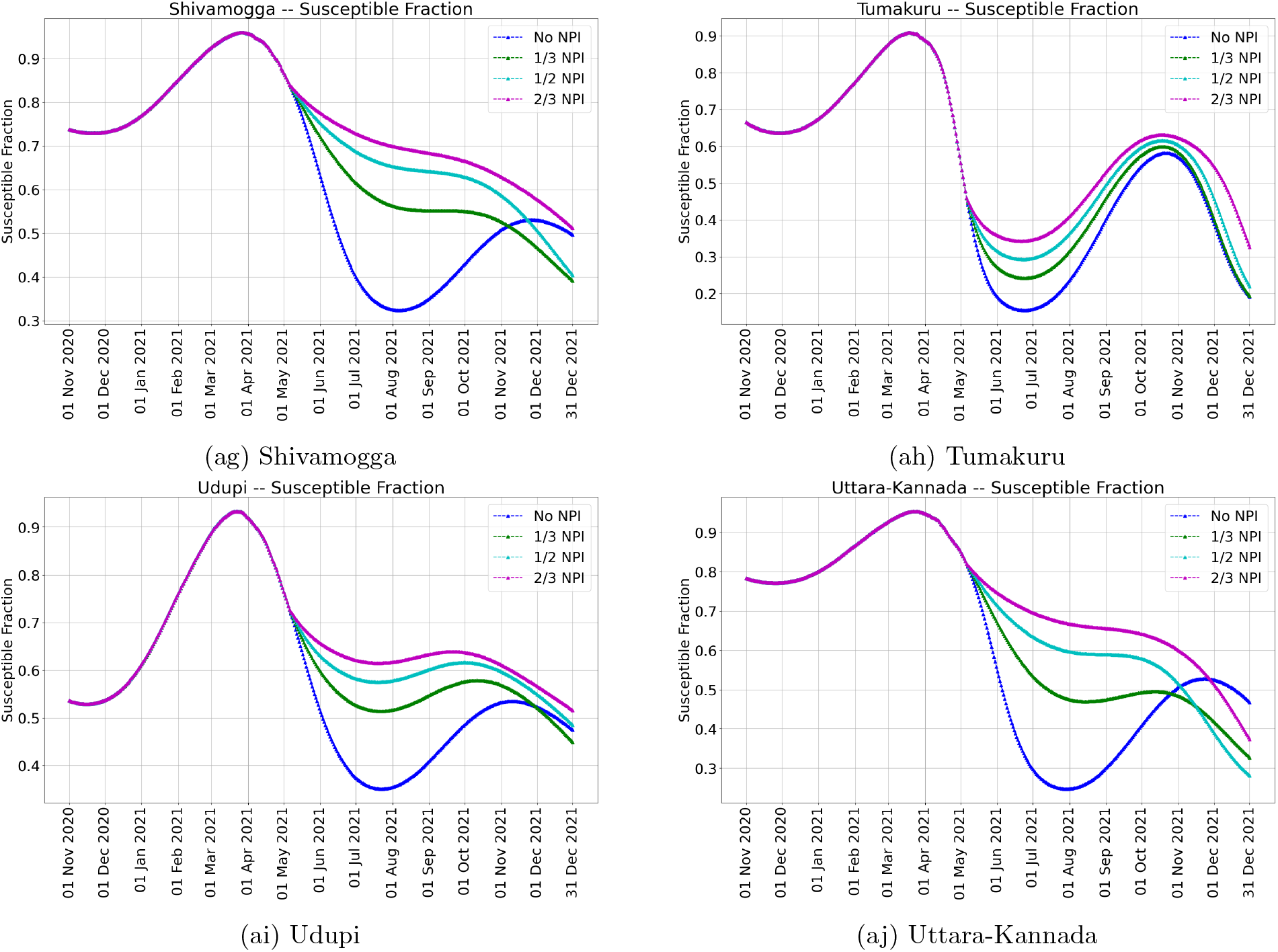
Susceptible fractions in each unit for various NPI intensities. Vaccination capacity is 167000 per day allocated in proportion to population in the units.

##### Validation Plots

**Figure 13:**
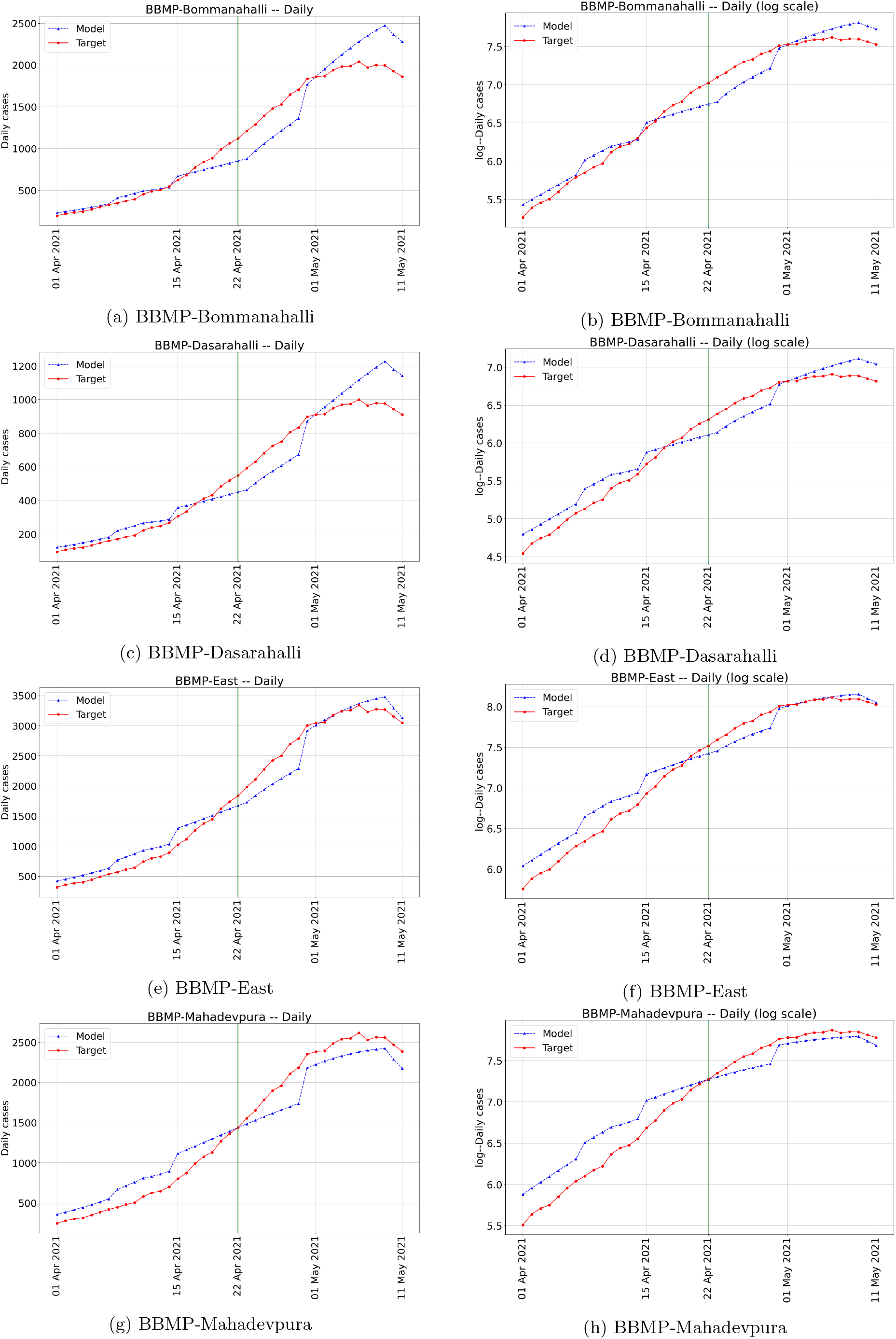
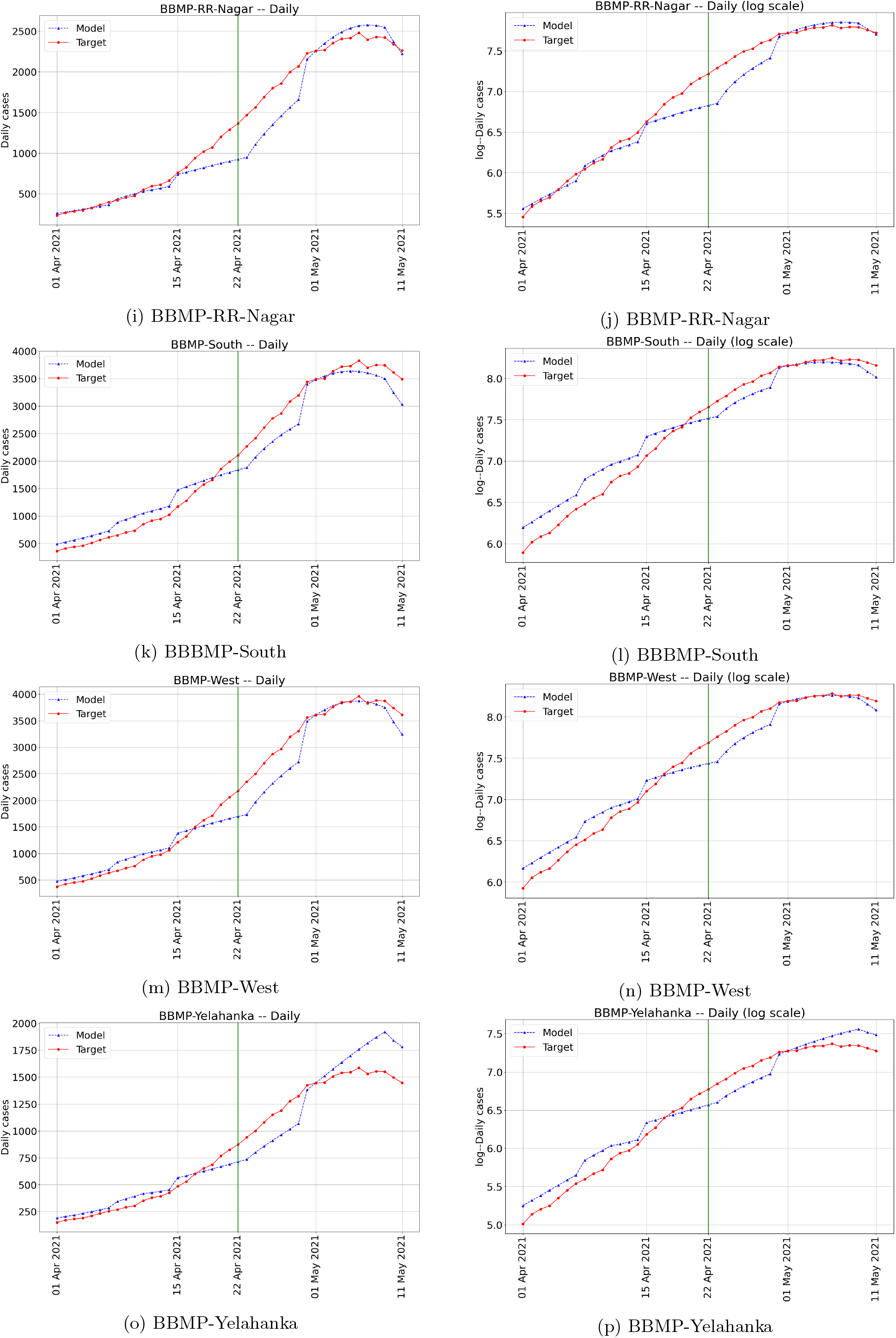
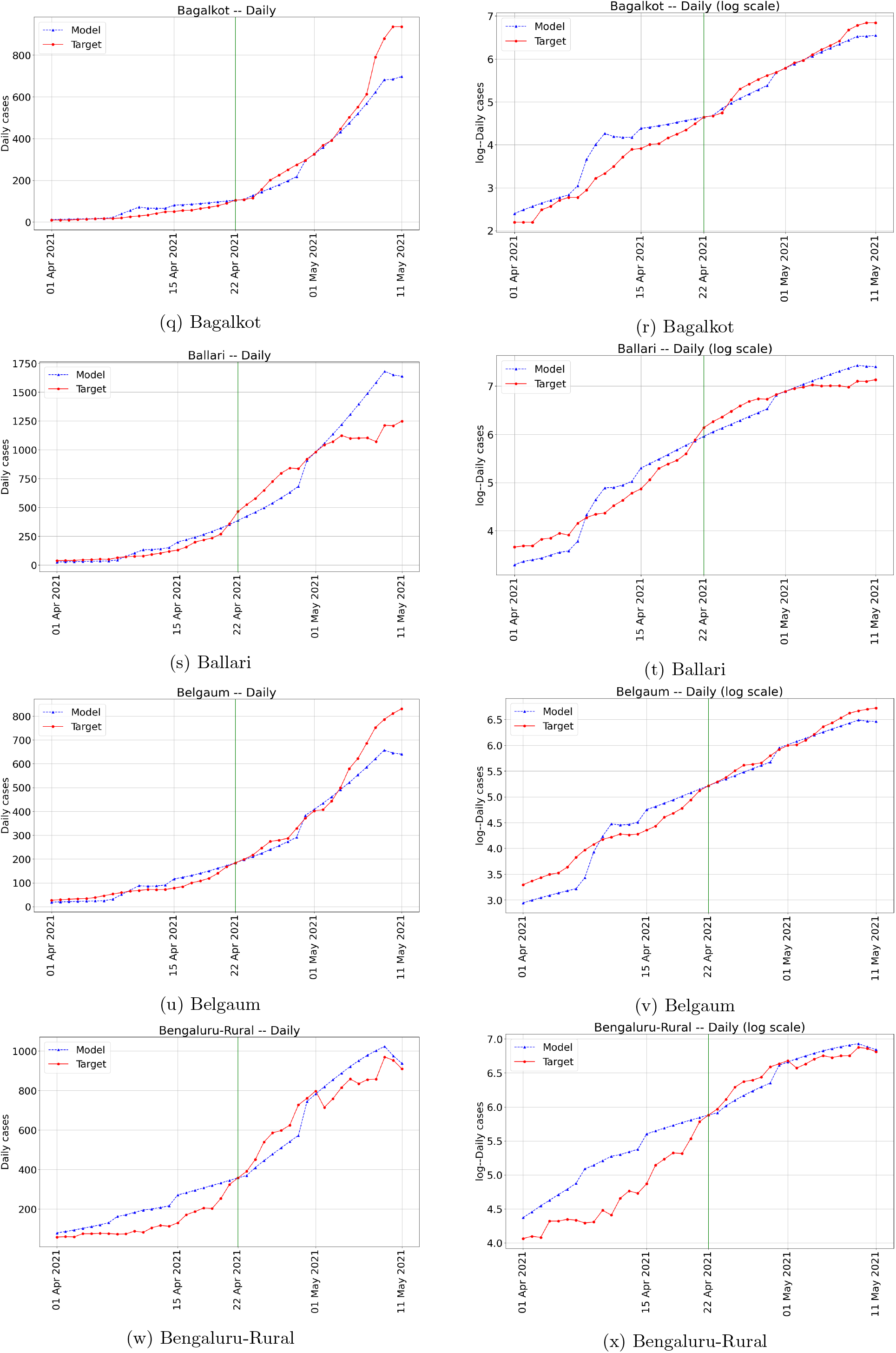
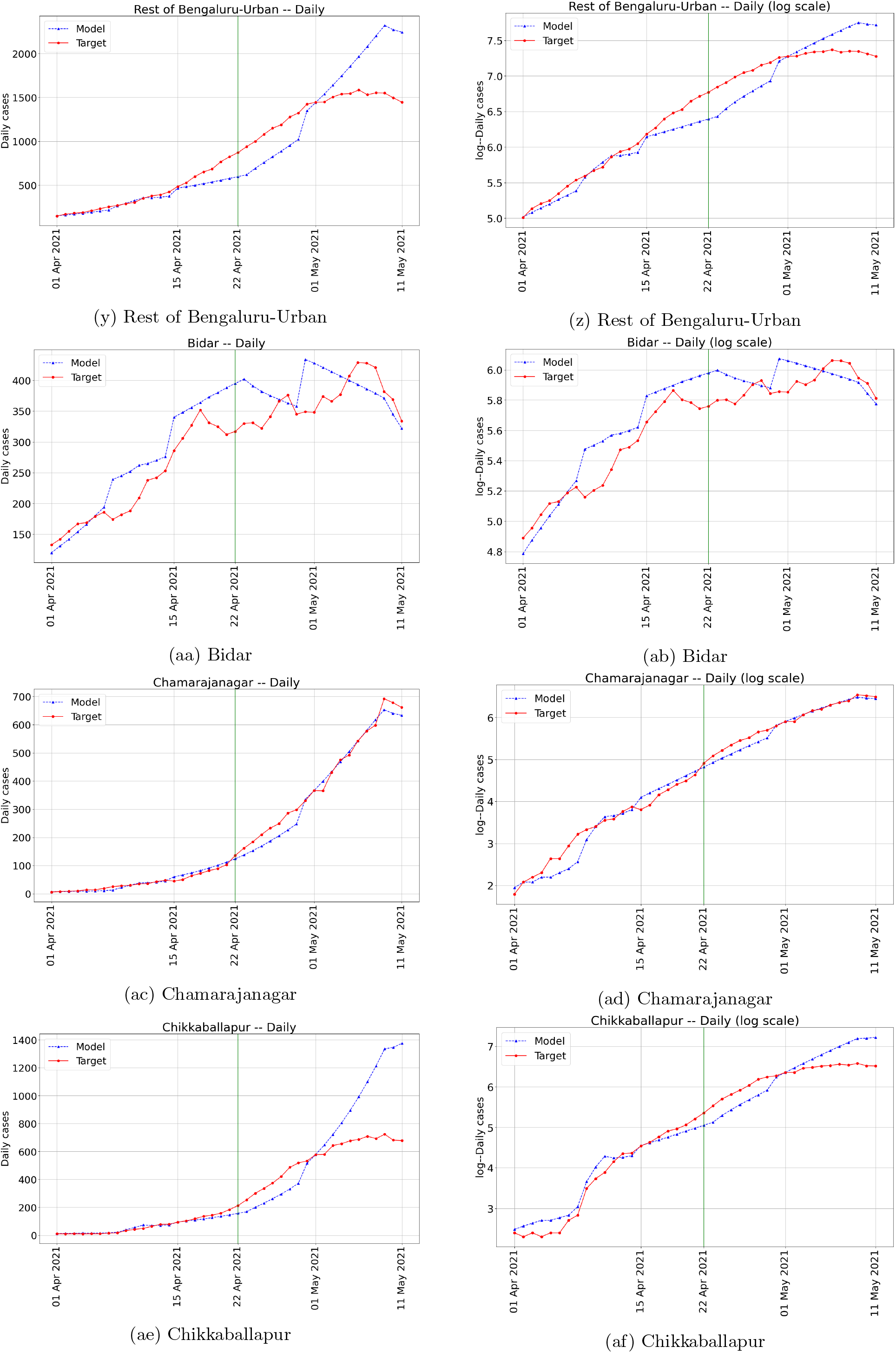
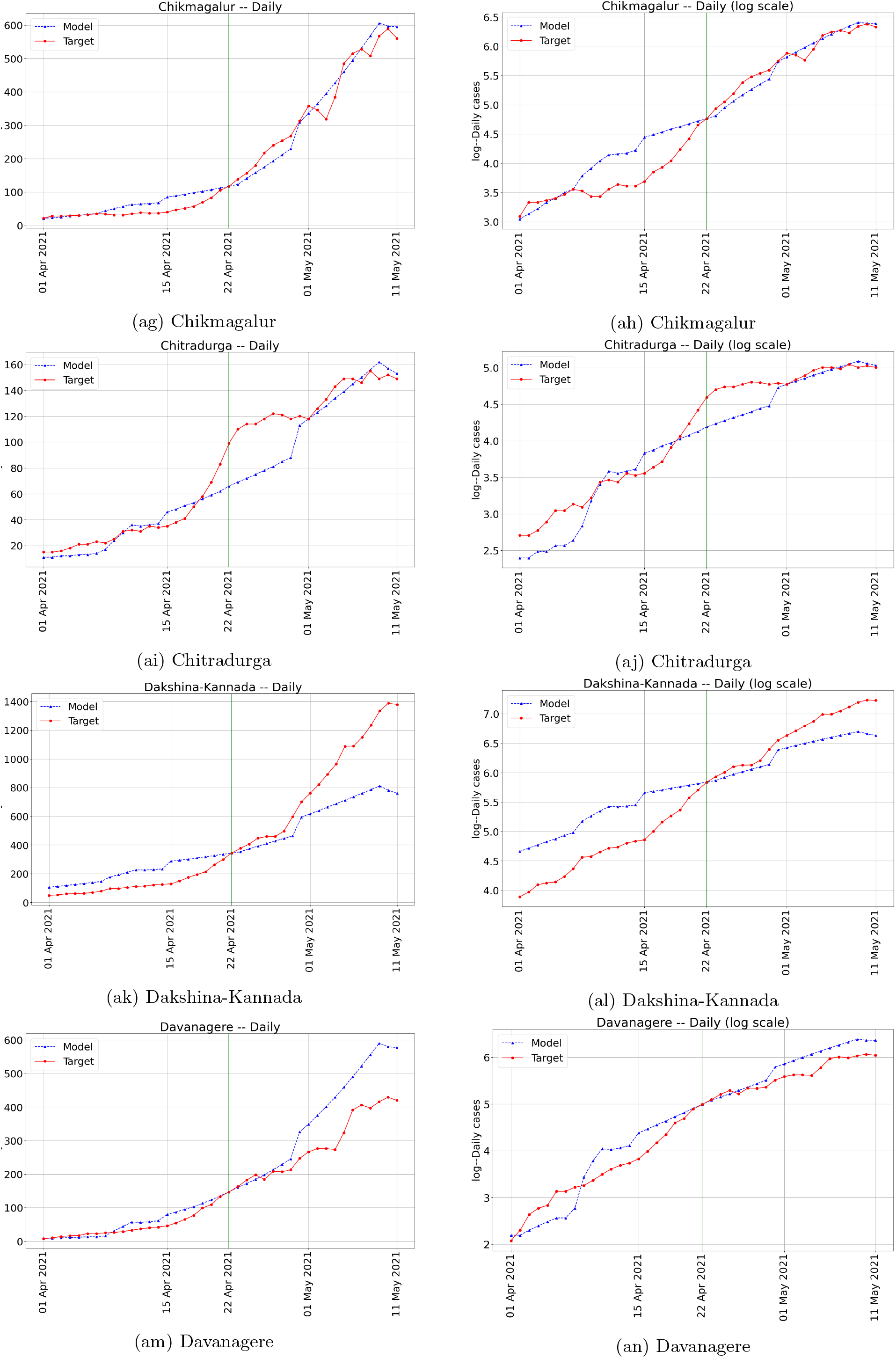
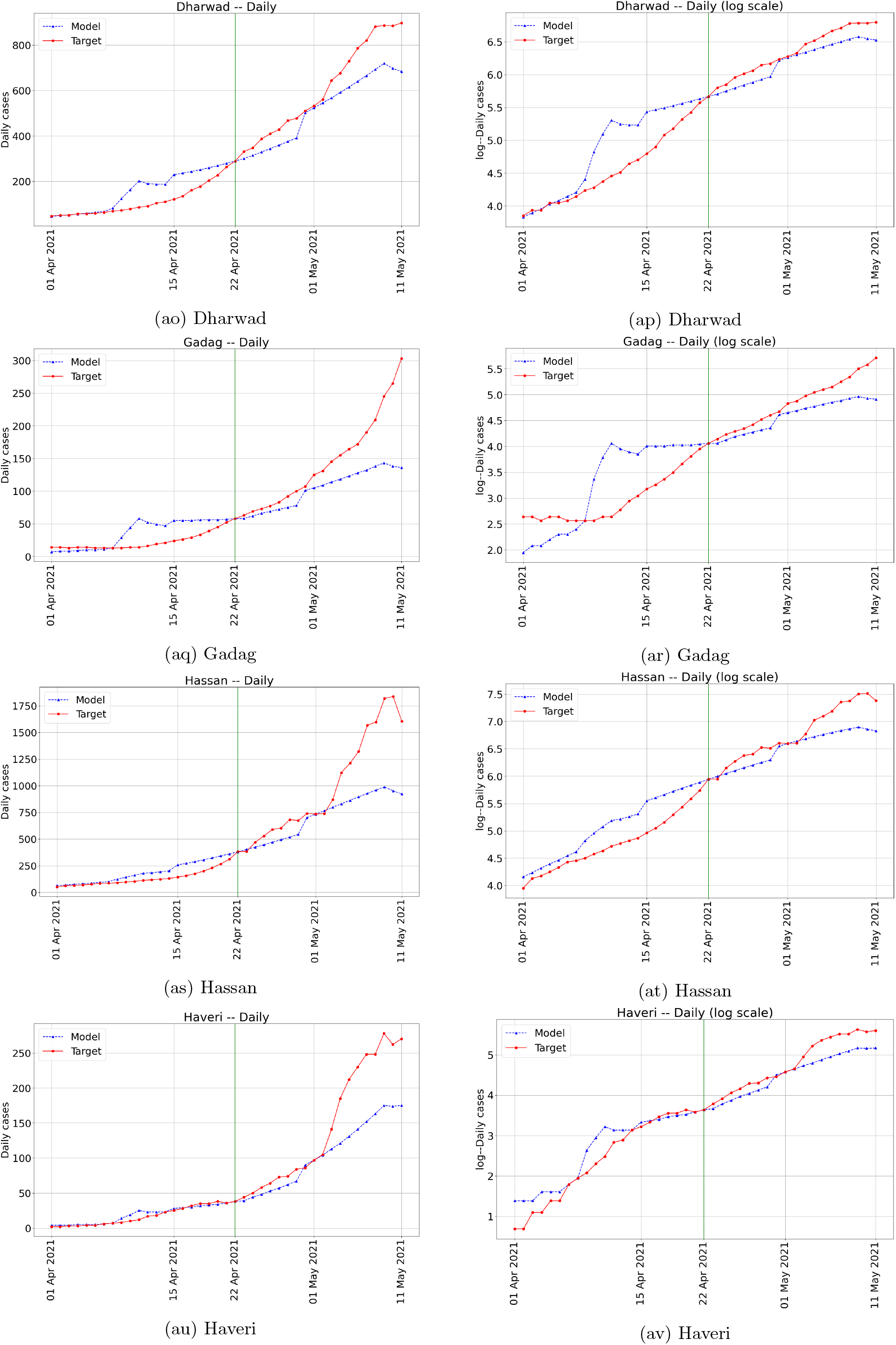
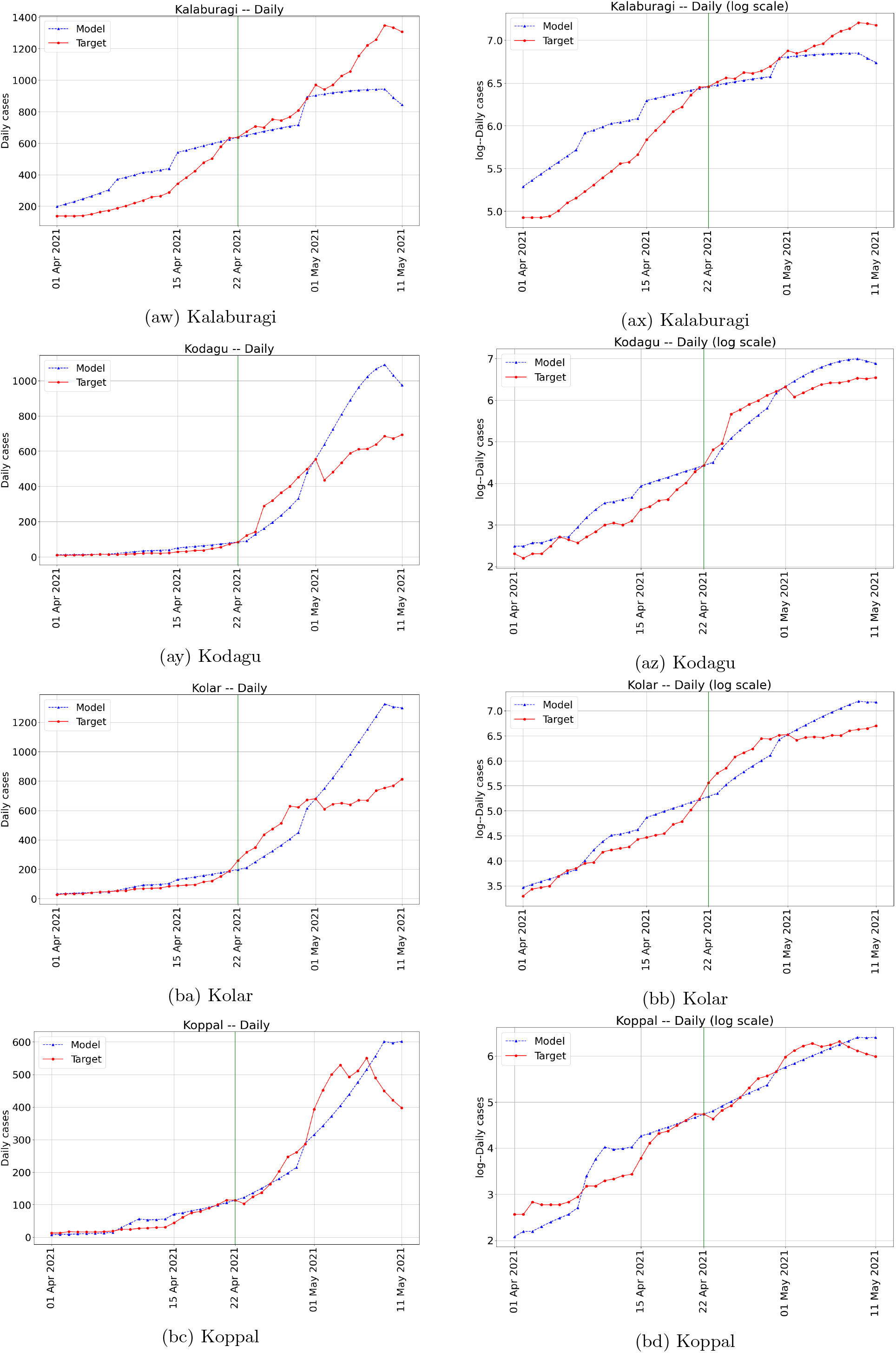
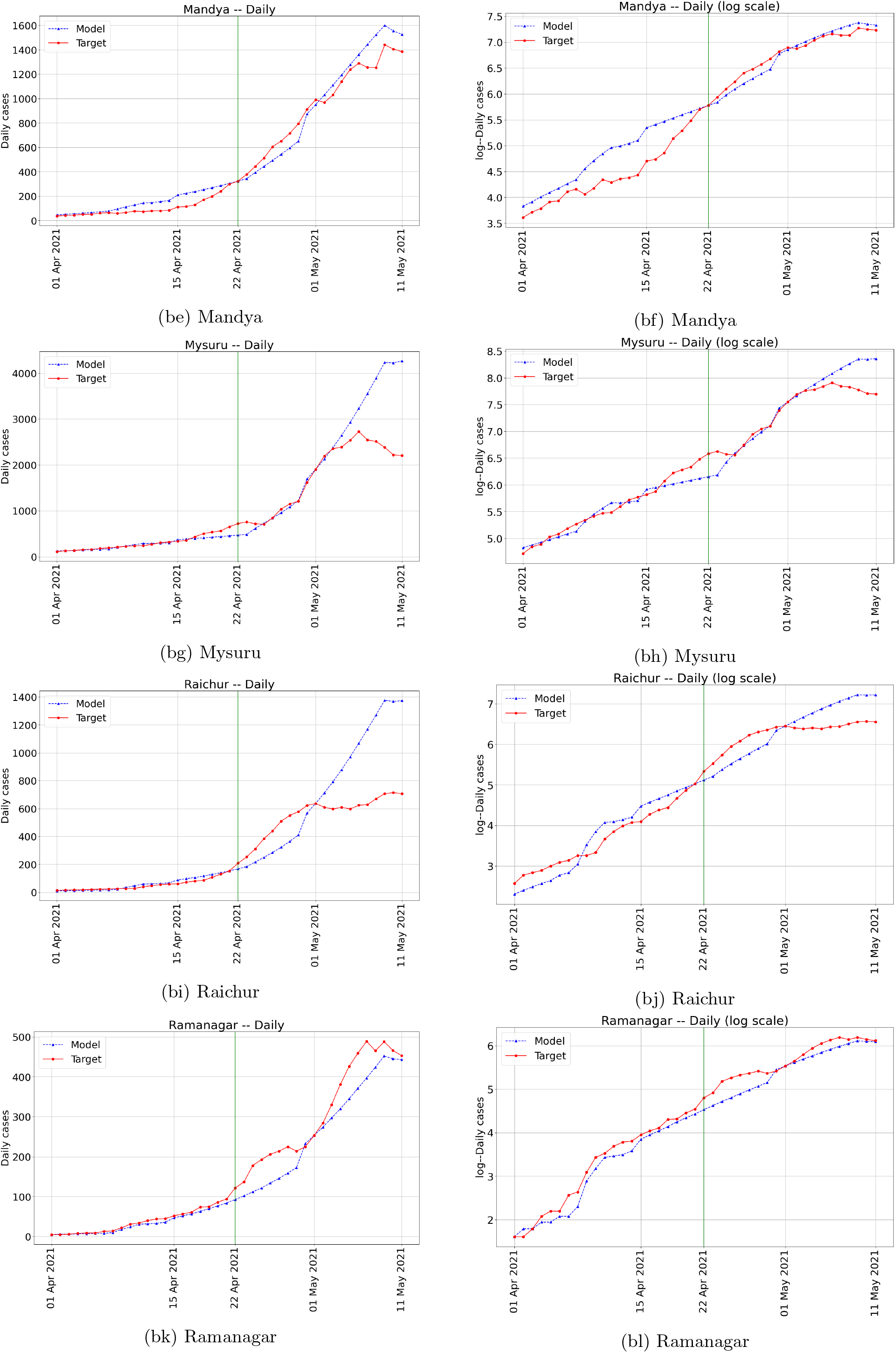
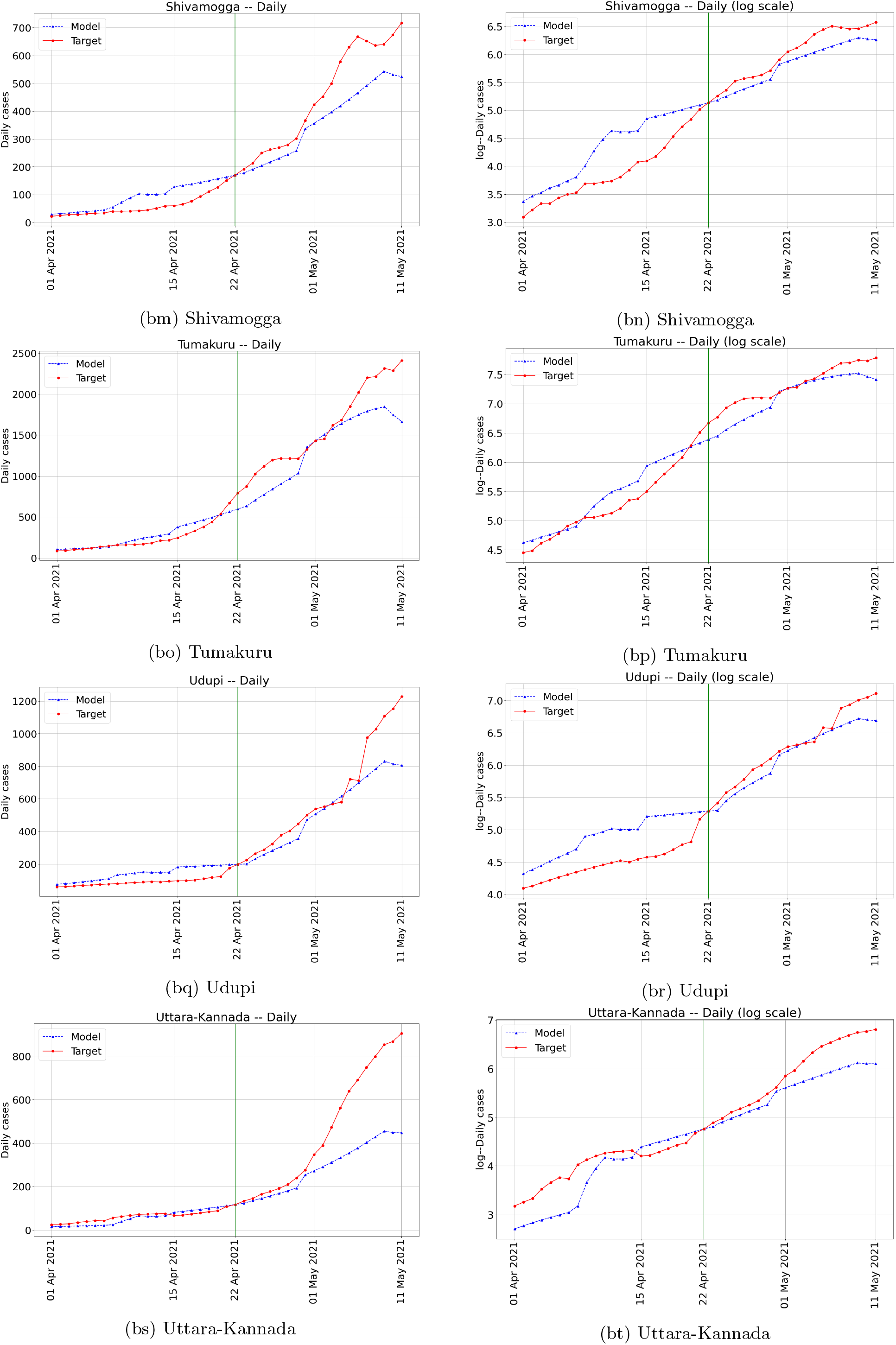
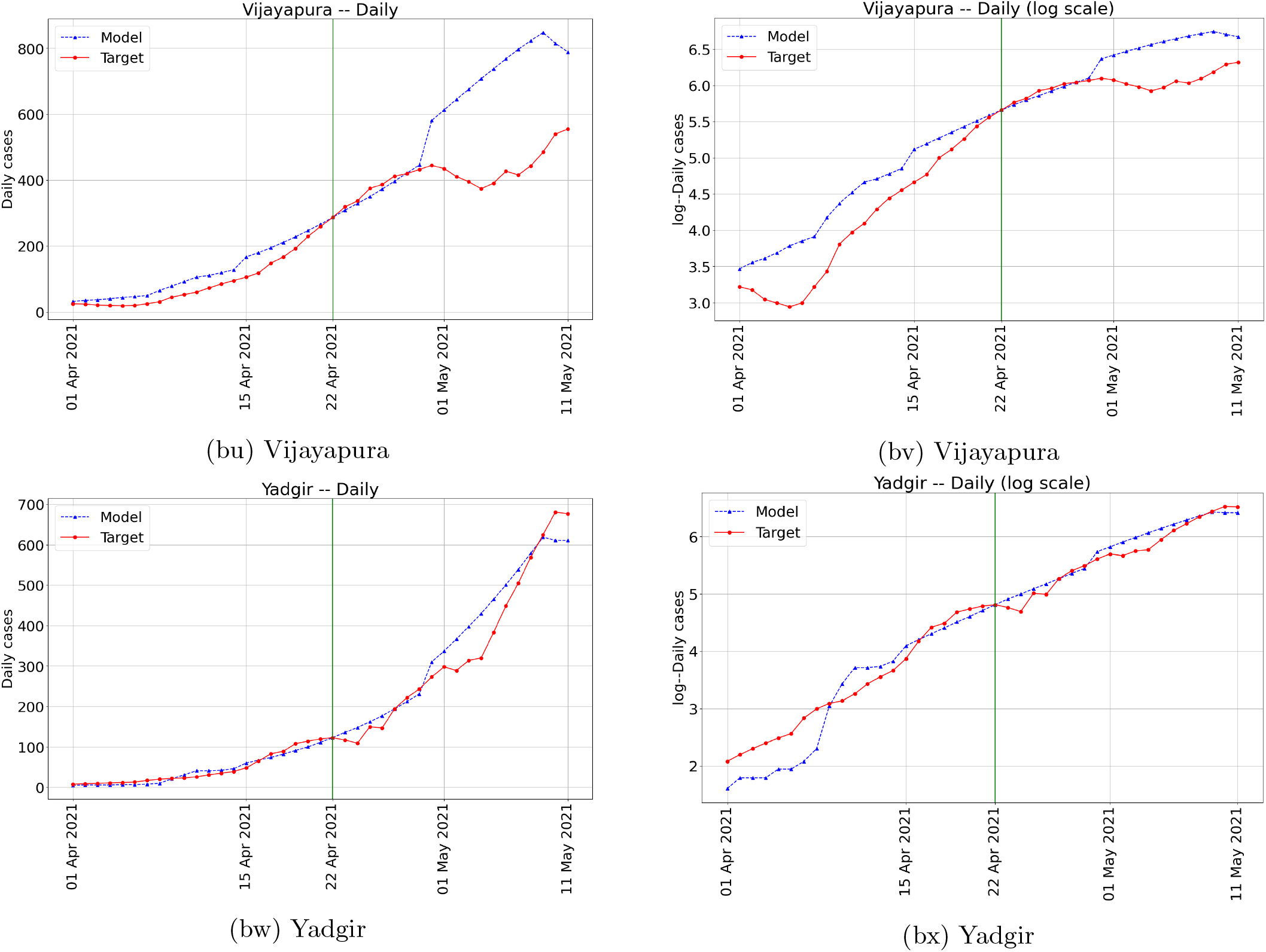
Validation plots for each unit. Calibration has been performed from 08–22 April 2021 and validated during 23 April – 11 May 2021. The vertical line separates the calibration and the validation phases.

## Footnotes

1 These include the closure of schools and colleges, workplaces, community spaces, mobility restrictions, and varying degrees of lockdowns.

2 But see [44] which is an Indian cohort study that reported only 85% seroconversion rate.

3 There has been a 50% increase in testing from 118,933 tests on 01 April 2021 [19] to 162,534 tests on 21 April 2021 [17].

4 At the time of writing, as indicated in the Introduction, two vaccines were being administered in India. The guidelines recommend two doses of the vaccine spaced twelve weeks apart. For effectiveness studies, see [47] and the interim phase 3 results [6].

5 To compare with the actual cumulative COVID-19 cases, one must add our increments from the reference date to the actual cumulative cases of 827,064 on 01 November 2020.

6 In the case of 2/3-NPI, the incidence rate proportional allocation does marginally better.

7 During the early exponential growth phase, the contact rate *β*(*t*) being static seems like a fair assumption. Further, during this early exponential growth phase, the susceptible fraction is likely quite large and the *s*(*t*) changes at a slower time-scale.

